# Dynamic Importance of Genomic and Clinical Risk for Coronary Artery Disease Over the Life Course

**DOI:** 10.1101/2023.11.03.23298055

**Authors:** Sarah M. Urbut, So Mi Jemma Cho, Kaavya Paruchuri, Buu Truong, Sara Haidermota, Gina Peloso, Whitney Hornsby, Anthony Philippakis, Akl C. Fahed, Pradeep Natarajan

## Abstract

**Importance:** Earlier identification of high coronary artery disease (CAD) risk individuals may enable more effective prevention strategies. However, existing 10-year risk frameworks are ineffective at earlier identification. Understanding the variable importance of genomic and clinical factors across life stages may significantly improve lifelong CAD event prediction.

**Objective:** To assess the time-varying significance of genomic and clinical risk factors in CAD risk estimation across various age groups.

**Design, Setting, and Participants:** A longitudinal study was performed using data from two cohort studies: the Framingham Offspring Study (FOS) with 3,588 participants aged 19-57 years and the UK Biobank (UKB) with 327,837 participants aged 40-70 years. A total of 134,765 and 3,831,734 person-time years were observed in FOS and UKB, respectively.

**Main Outcomes and Measures:** Hazard ratios (HR) for CAD were calculated for polygenic risk scores (PRS) and clinical risk factors at each age of enrollment. The relative importance of PRS and Pooled Cohort Equations (PCE) in predicting CAD events was also evaluated by age groups.

**Results:** The importance of CAD PRS diminished over the life course, with an HR of 3.58 (95% CI 1.39-9.19) at age 19 in FOS and an HR of 1.51 (95% CI 1.48-1.54) by age 70 in UKB. Clinical risk factors exhibited similar age-dependent trends. PRS significantly outperformed PCE in identifying subsequent CAD events in the 40-45-year age group, with 3.2-fold more appropriately identified events. The mean age of CAD events occurred 1.8 years earlier for those at high genomic risk but 9.6 years later for those at high clinical risk (p<0.001). Overall, adding PRS improved the area under the receiving operating curve of the PCE by an average of +5.1% (95% CI 4.9-5.2%) across all age groups; among individuals <55 years, PRS augmented the AUC-ROC of the PCE by 6.5% (95% CI 5.5-7.5%, p<0.001).

**Conclusions and Relevance:** Genomic and clinical risk factors for CAD display time-varying importance across the lifespan. The study underscores the added value of CAD PRS, particularly among individuals younger than 55 years, for enhancing early risk prediction and prevention strategies.

**Key Points Question:** How do genomic and clinical risk factors contribute to coronary artery disease (CAD) risk across a broad age range?

**Findings:** This longitudinal observational study across two cohorts found that both genomic and clinical risk factors exhibit age-dependent significance for CAD risk. Polygenic risk scores (PRS) are most informative for individuals younger than 55 years, improving the predictive accuracy of current risk equations for these individuals.

**Meaning:** The study emphasizes the need to incorporate the dynamic effects of cardiovascular risk factors, particularly genomic risk, for more accurate early-life risk prediction and efficient CAD prevention strategies.

## Introduction

Accurate risk estimation for coronary artery disease (CAD) early in the life course is a major goal in medicine, as CAD remains the leading cause of mortality and morbidity.^1^ Since coronary atherosclerosis often begins early in life and progresses over the life course, early identification of high-risk individuals offers the possibility for substantial risk mitigation.^2^

There are several reasons why contemporary risk estimators in clinical practice do not adequately identify high-risk individuals early in life. First, guideline-based risk calculators are valid only for ages 40 years or older and are often limited to short-term (e.g. 10-year) fixed-time horizons.^3,4^ Therefore, chronologic age remains the primary determinant of estimated 10-year risk, and high risk cannot be identified earlier in life, thereby delaying effective prevention opportunities.^5^ Second, even when prediction is extended to estimate lifetime risk, it fails to capture the dynamic trajectory of an individual’s changing risk profile, such as changing biomarker, biometric measurements, or lifestyle. Finally, models are developed assuming proportional hazards, which impose that the effect of each risk factor is either constant over the baseline hazard ratio through life or that interaction is a linear function of time. Both assumptions are empirically inaccurate for CAD clinical risk factors.^6^

CAD polygenic risk score (PRS) has emerged as a tool to estimate risk complementary to clinical risk factors and is uniquely available very early in life. Traditional models considering these scores often employ a fixed-time horizon and rely on assumptions that do not hold true for the dynamic and complex landscape of CAD risk factors. We aim to break down existing barriers in CAD risk prediction by integrating both genomic and clinical risk factors in a single, dynamically-adjusting model. Using two cohorts ranging from 19 to 70 years of age, followed for up to 44 years, we illuminate how the relevance of these risk factors shifts over an individual’s life course, thereby offering a more nuanced and applicable framework for CAD risk estimation. While recent work by Marston et al. in the UK Biobank has shown that CAD PRS carries greater effects for younger people,^7^ its comparative and complementary performance with clinical risk calculators is less clear for both premature and cumulative events across a broad age range. The integration of genomic and clinical risk in a single model continues to be a barrier to clinical implementation of CAD PRS at scale. Such integration will ideally incorporate the dynamic importance of genomic and clinical risk for CAD over the life course for optimal utility.

Here, we leverage two cohorts of individuals enrolled across the ages of 19 to 70 years and followed for up to 44 years to show that genomic and clinical risk factors vary in their importance over the life course and to explain a changing proportion of variation for CAD risk. We show that CAD PRS adds the most information for young and early middle-aged individuals when compared with older individuals and predicts a greater number of both premature and overall events for younger individuals. This framework mitigates current age-dependent limitations of CAD clinical risk scores.

## Methods

### Study participants

Two cohorts were included in this study. First, the Framingham Offspring Study (FOS) is a longitudinal US-based cohort study consisting of the children of the original participants of the Framingham Heart Study, recruited between 1971 and 1975 and followed through 2018.^9^ Clinical data on cardiovascular risk factors and incident disease were available for 3,821 participants, and genetic data for a subset (N=2,754), through the database of Genotypes and Phenotypes (dbGaP; accession phs000007.v33.p14). We conducted an analysis of clinical risk factors on the complete dataset (N=3,588) and the genetic analysis on the subset (N=2,629), after excluding 233 individuals for missing risk factor data, current lipid-lowering medication, or pre-existing CAD (Supplementary Figure 1).

Second, the UK Biobank (UKB) is a prospective nationwide population-based study that enrolled middle-aged adults between 2006 and 2010 and followed through present.Examiners collected baseline phenotypic, genetic, self-reported, and electronic health records on 502,485 participants.^10^ In the present study, we included 327,837 participants from the UKB after excluding 174,378 who lacked quality-controlled genotyping, risk factor, lipid, or medication information or carried a diagnosis of CAD at baseline (Supplementary Figure 2).

Informed consent was obtained from all participants, and secondary data analyses of dbGAP based FOS and UKB were approved by the Mass General Brigham Institutional Review Board applications 2016P002395 and 2021P002228.

### Study outcomes

In the FOS, CAD was defined as coronary death or myocardial infarction and recorded by independent reviewers over a follow-up period of a median 43.0 [Interquartile Range (IQR) 38.6-47.4] years encompassing 134,765.2 person-years, using medical histories, physical examinations at the study clinic, hospitalization records, and communication with participants’ physicians, as previously described.^11,12^

In the UKB, participants were followed for a median of 12.2 [IQR 11.4-15.1] years encompassing 3,831,734 person-years. CAD was defined as a composite of myocardial infarction, coronary revascularization, or death related to either as previously described.^13^ Myocardial infarction was based on self-report or hospital admission diagnosis as performed centrally and recorded in I21-I21.4, I21.9, I22-I22.1, I22.8, I22.9, I23-I23.6, I23.8, I24-I24.1, I24.8,I24.9, I25.2.^13^ Coronary revascularization was assessed based on an OPCS-4 coded procedure for coronary artery bypass grafting (K40.1–40.4, K41.1–41.4, K45.1–45.5) or coronary angioplasty with or without stenting (K49.1–49.2, K49.8–49.9, K50.2, K75.175.4, K75.8–75.9).

### Genomic risk

CAD PRS, a measure of the cumulative risk from many genetic variations across the genome, was used to quantify genomic risk.^14^ Genetic data for the FOS were made available from the NHLBI SNP Health Association Resource (SHARe) project, in which genotyping was conducted using approximately 550,000 SNPs (Affymetrix 500K mapping array plus Affymetrix 50K supplemental array) and imputed using the 1000 Genomes reference panel as reported previously.^9^ The genetic data for UKB was phased and imputed centrally to ∼96 million variants with the Haplotype Reference Consortium (HRC) and the UK10K + 1000 Genomes reference panel.^10^ In both cohorts, we computed a CAD PRS using publicly available weights for GPS_CAD_, a genome-wide polygenic score for CAD consisting of 6.6 million variants.^14^ In clarifying analyses, participants were classified as having low genomic risk if they fell in the bottom quintile, intermediate genomic risk if they fell in the middle three quintiles, and high genomic risk if they fell in the top quintile, of the population distribution of PRS.

### Clinical risk factors

Individual clinical risk factors of CAD as well as a guideline-supported clinical risk score (i.e., the Pooled Cohort Equations [PCE]) were used to estimate CAD risk. Clinical risk factors such as current smoking, diagnosis of diabetes, antihypertensives prescription, blood pressure, and lipids were collected at cohort enrollment based on a combination of self-report, blood test, and medical chart review.^11^ Systolic blood pressure measurement was adjusted for anti-hypertensive medication use by adding 15mmHg. Lipids were adjusted for the use of lipid-lowering medication by dividing the LDL-C and total cholesterol value by 0.7 and 0.8, respectively, as previously described.^15^

The PCE was computed in the UKB, which provided a ten-year risk estimate of atherosclerotic cardiovascular disease (ASCVD).^16^ Guideline-based risk strata were indicated as follows: low or borderline (<7.5%), intermediate (≥7.5 to <20%), and high (≥20%).^16^

### Statistical Analysis

At enrollment, we computed the age-specific hazard ratios (HRs) and proportions of variation explained by each risk factor for cumulative risk of CAD. We divided each dataset into individuals whose age at enrollment and baseline ascertainment of risk factor levels were within one calendar year of each age under consideration. We report the results from a locally estimated smoothed scatter (loess)^17^ weighted according to the tricube distance function to borrow information from nearby windows. After confirming that the Cox proportional hazard assumption was now satisfied by this approach (Supplementary Figure 3, Supplementary Methods), we reported the average HR and proportion of variation explained (PVE) of CAD over the study period with respect to one unit increase in standardized risk factor for individuals within one calendar year of assessment (Supplementary Methods).

For age-dependent relative incidence analyses, we computed the incidence rates for each CAD PRS percentile and divided by the incidence rate for those individuals of the lowest risk percentile per age group, so that the lowest age-relative incidence rate equals one. For cumulative hazard analyses, we computed cumulative hazard in strata of PRS and PCE within each age category (younger than 55 years, 55-65 years, and older than 65 years). Within each age category, we then stratified by PRS category (bottom quintile, middle three quintiles, top quintile) and then by age-specific PCE risk categories (bottom quintile, middle three quintiles, top quintile).

For prediction of cumulative events, we identified individuals with a diagnosis of CAD over the observed time-period and computed the number of events that were predicted for individuals categorized as intermediate or high risk by PCE (10-year ASCVD risk ≥7.5%), high polygenic score (top quintile) at age of enrollment, or both. Traditional area-under-the-curve (AUC) was evaluated for development of CAD on PRS or PCE categories based on logistic regression and fitted for each age group separately.

## Results

### Study Participants

We studied two cohorts free of cardiovascular disease at baseline and spanning the life course: (i) FOS comprising 3,588 individuals (50.9% female) ages 19-50 years at enrollment and followed for a median of 43.7 (interquartile range [IQR] 38.7-47.4) years and (ii) UKB, comprising 327,837 participants (57% female) ages 40-70 years at enrollment followed for a median of 12.1 (IQR 11.4-12.7) years (Table 1). Apart from smoking, clinical risk factors were more prevalent in the UKB as expected given the age differences. For example, 1581 (44%) of FOS participants (enrolled 1971-1975) were current smokers, compared to 33,869 (10%) of UKB participants (enrolled 2006-2010). During follow-up, 695 (19.4%) of FOS participants and 11,190 (3.4%) of UKB participants developed CAD. Of those incident events, the proportion of premature CAD events – defined as occurring before age 55 years – were 179 of 695 (25.8%) in the FOS and 1085 of 11,190 (9.7%) in the UKB, respectively.

**Table 1.** Characteristics of Study Participants from the FOS (N=3,588) and UKB (N=327,837)

| **Characteristics** | **FOS (N=3,588)** | **UKB (N=327,837)** |
| --- | --- | --- |
| Age at risk estimation, mean (SD), years | 35.9 (10.2) | 56.1 (8.1) |
| Female, n (%) | 1,828 (50.9) | 186,507 (56.9) |
| White race, n (%) | 3,588 (100) | 274,927 (83.9) |
| Incident CAD, n (%) | 695 (19.3) | 11,190 (3.4) |
| Follow-up period, median [IQR) | 43.7 [38.7-45.3) | 12.1 [11.4-12.7) |
| Diabetes mellitus, n (%) | 27 (0.7) | 2413 (0.7) |
| Current smoking, n (%) | 1581 (44.1) | 33869 (10.3) |
| Total cholesterol, mean (SD), mg/dL | 197 (38.8) | 228.6 (41.4) |
| HDL Cholesterol, mean (SD), mg/dL | 52.1 (16.0) | 57.2 (14.8) |
| LDL cholesterol, mean (SD), mg/dL | 127 (36.6) | 144.0 (31.9) |
| Triglycerides, mean (SD), mg/dL | 99.1 (86.7) | 151.9 (90.3) |
| Diastolic blood pressure, mean (SD), mmHg | 78.5 (10.9) | 82.8 (11.2) |
| Systolic blood pressure, mean (SD), mmHg | 121 (16.4) | 139.7 (20.4) |
| Taking antihypertensive medication, n (%) | 102 (2.8) | 41,088 (12.5) |
| PCE 10-year risk category |  |  |
| Low or borderline (<7.5%), n (%) | - | 207,150 (63.2) |
| Intermediate (≥7.5 to <20%), n (%) | - | 96,775 (29.5) |
| High (≥20%), n (%) | - | 23,912 (7.3) |
| Genetic data available, n (%) | 2,656 (72.5) | 327,837 (100.0) |
| CAD polygenic risk score category |  |  |
| Low, n (%) | 531 (20.0) | 65,696 (20.0) |
| Intermediate, n (%) | 1,593 (60.0) | 196,750 (60.0) |
| High, n (%) | 532 (20.0) | 65,391 (20.0) |

Characteristics for study participants from the Framingham Offspring Study (FOS) and UK Biobank are reported for all individuals based on data obtained at enrollment. CAD: coronary artery disease, PRS: polygenic risk score, PCE: Pooled Cohort Equations, HDL: High-density lipoprotein cholesterol, LDL: Low-density lipoprotein cholesterol.

### Age-dependent effects of genomic and clinical risk factors

We calculated the hazard ratio of CAD per standard deviation of PRS at each age of enrollment. The HR per standard deviation of CAD PRS decreased over the life course – from 3.58 (95% CI 1.39-9.19) at age 19 years to 1.99 (95% CI 1.06-3.70) at age 56 years in FOS, and from 2.25 (95% CI 1.77-2.87) at age 41 years to 1.39 (95% CI 1.30-1.48) by age 70 years in UKB (Figure 1, Supplementary Tables 1 and 2).

**Figure 1.**
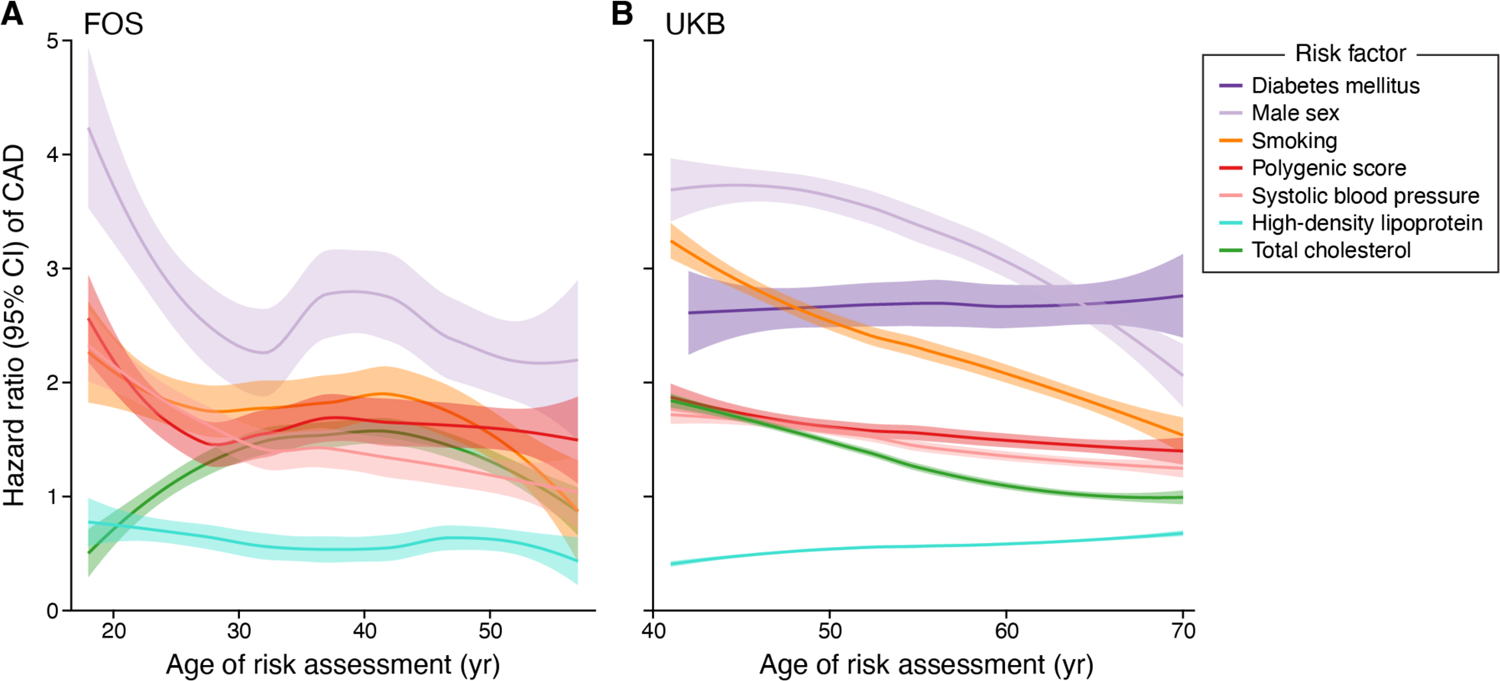
Dynamic Hazard Ratio of CAD for Genomic and Clinical Risk Factors by Age at Estimation The age-specific hazard ratio (HR) for risk of CAD is plotted for multiple risk factors at each age of enrollment (A) between 19 and 57 years in the FOS (N=3,588), and (B) between 40 and 70 years in the UK Biobank (N=327,837). The HR is obtained from Cox proportional hazards estimate at each age of enrollment for a standardized unit increase in each of polygenic score, total cholesterol, HDL cholesterol, and systolic blood pressure or a binary indicator for smoking, male sex, and diabetes mellitus (only in the UK Biobank given the low prevalence of diabetes mellitus in FOS). We note the different time periods (1970-2010 and 2007-2021) for FOS and UKB, respectively. The difference in absolute values can thus not be considered as levels of these clinical risk factors varied between populations (Table 1) and between eras. Accordingly, excluding individuals on statins results in a different population in each cohort. No covariates are used in the analysis to isolate the effect of each risk factor separately. **HR:** Hazard Ratio**; HDL**: High density lipoprotein, **CAD**: coronary artery disease, **FOS**: Framingham Offspring Cohort, **95% CI:** 95% Confidence Interval.

We next calculated the HR of clinical risk factors at each age of enrollment and similarly observed decreasing hazard ratios over the life course. For example, the HR (95% CI) of CAD for smoking decreased from 1.98 (0.44-8.84) at age 19 years to 0.98 (0.41-2.33) at age 56 years in the FOS and from 3.51 (2.13-5.80) at age 41 years to 1.62 (1.28-2.04) at age 70 years in the UKB. The trends were similar for systolic blood pressure and diabetes (Figure 1, Supplementary Tables 1 and 2). Excess risk associated with male sex similarly declined with age – from 3.29 (95% CI 0.64-16.95) at age 19 to 2.59 (95% CI 0.92-7.25) at age 57 in the FOS and from 3.20 (95% CI 1.82-5.64) at age 41 to 1.99 (95% CI 1.74-2.26) at age 70 in the UKB (Figure 1, Supplementary Tables 1 and 2).

When clinical risk factors were considered in composite as part of the PCE, the HR for CAD for a 1% increase in estimated 10-year risk remained relatively stable over the life course – 1.24 (95% CI 1.18-1.30) at age 41 years and 1.04 (95% CI 1.03-1.04) at age 70 years (Supplementary Figure 4). However, when scaling the PCE by its SD of 7.2%, HR (95% CI) per SD ranges from 4.4 (3.31-5.95) at age 41 years to 1.3 (1.29-1.31) at age 70 years (Supplementary Figure 4). A high PCE was exceedingly rare among young participants (0.14%, 95% CI 0.13-0.16) (Supplementary Figure 5).

We next computed the PVE of CAD on each risk factor for individuals up to and including the age in question. We observed a decreasing PVE with increasing age for PRS, from 19% (95% 18.9-19.1) at age 19 years to 3.2% (95% CI 3.19-3.21) at age 57 years in the FOS and from 5.9% (95% CI 5.89-5.91) at age 40 years to 1.7% (95% CI 1.69-1.71) at age 70 years. (Supplementary Figure 4, Supplementary Tables 3 and 4).

### Relative importance of genomic and clinical risk of CAD by age

To compare the relative importance of genomic versus clinical risk, we limited our analysis to the UKB where both could be calculated. The distributions of PRS of participants across all age groups were similar and the absolute risk of CAD increased with increasing PRS (Figures 2A and 2B, Supplementary Figure 6). Over the study period, the absolute CAD risk difference between those < 55 years in the 1^st^ and 99^th^ percentiles was 3.1%, while at >65 years rose to 7.1% (Figure 2A). However, the corresponding relative risks were 5.2-fold (95% CI 5.1-5.4) and 3.2-fold (95% CI 3.1-3.3), respectively (Figure 2C).

**Figure 2.**
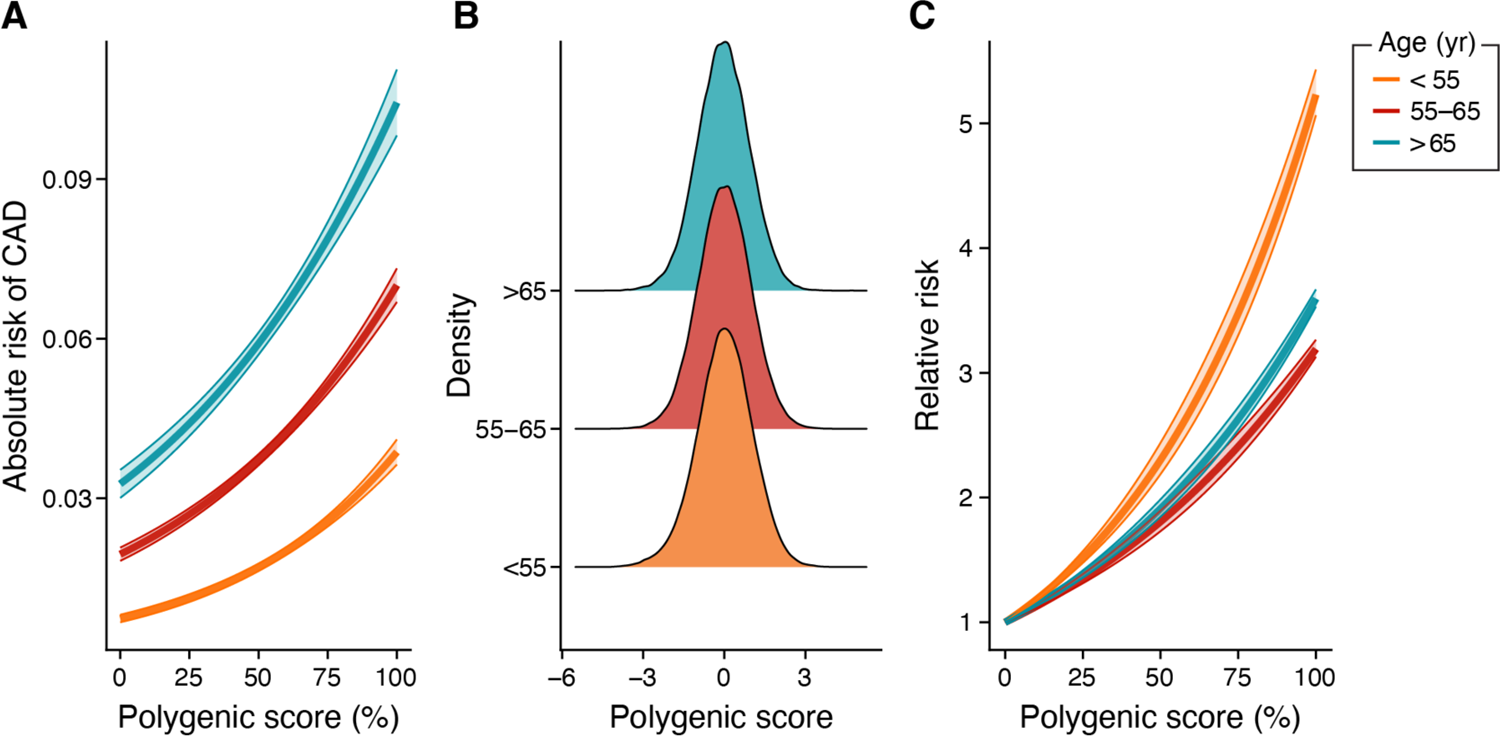
Absolute and Relative Incidence Rate of CAD by Genomic Risk per Age Group In the UK Biobank (N=327,837), three age groups (<55, 55-65, and >65 years) at risk estimation were used to compare the stratification of the observed absolute and relative risk across polygenic score percentile. (A) The absolute risk of CAD increased with increasing polygenic score percentile in all three age groups, and older participants had higher absolute risk of CAD. Absolute risk of CAD ranged from 0.7 to 3.9% in the <55 years age group, 1.9 % to 7.0 % in the 55-65 years age group, and 3.3 to 10.4% in the >65 years age group. (B) The polygenic score distribution was similar across three age groups. (C) Relative risk gradient of genomic risk is greatest for younger age groups. The 99th percentile of polygenic score was associated with a 5.2-fold increase in risk in the <55 years age group, 3.6-fold increased risk in the 55-65 years age group, and 3.2-fold increase in risk in the >65 years age group. **CAD**: coronary artery disease. **PRS:** Polygenic risk score.

When classifying PCE and PRS strata within each age group as high (top quintile), intermediate (middle three quintiles), and low (bottom quintile) (Supplementary Table 5), there was a marked gradient of cumulative hazard of CAD events over the 12-year follow-up period (Figure 3). This stratification was highest in the <55 years age group, ranging from 0.045% (95% CI 0.23-0.67) for individuals with low PRS and low PCE to 14.6% (95% CI 12.8-15.5) for individuals with high PRS and high PCE. The corresponding stratification in the >65 years age group was 4.6% (95% CI 0.01-0.09) to 37.6% (95% CI 0.11-0.64) (Figure 3).

**Figure 3.**
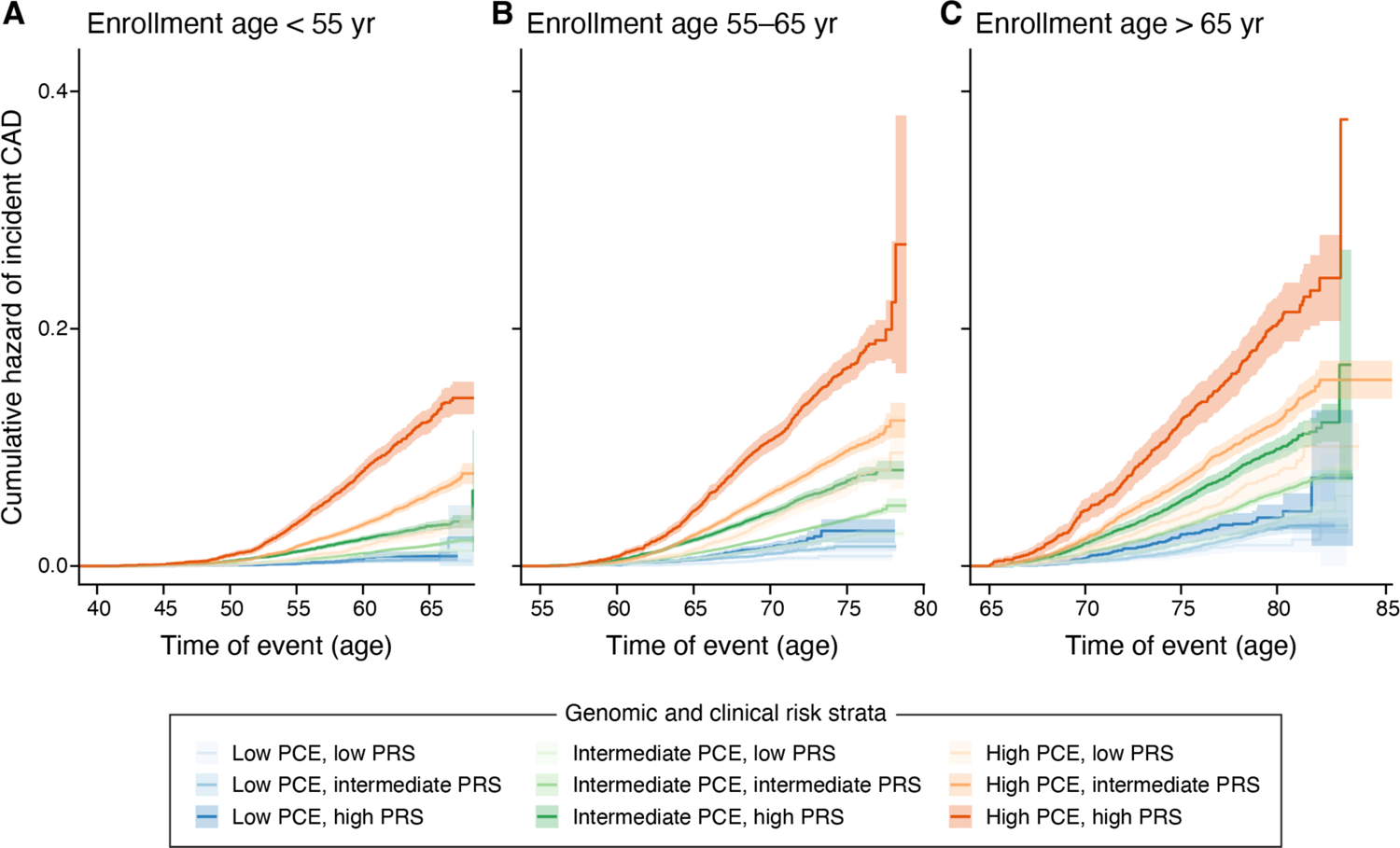
Cumulative Hazard of Incident CAD by Clinical and Genomic Risk in Three Age Groups In the UK Biobank (N=327,837), three age groups (<55, 55-65 and over 65 years) at risk estimation were used to compare the cumulative incidence of CAD by genomic (PRS) and clinical (PCE) risk levels defined as low (bottom quintile), intermediate (middle three quintiles), and high (top quintile) within each age group. We report the cumulative hazard over the observed follow-up time (median 12.2 years). The stratification was highest in the <55 years age group (A), where the cumulative hazard ranged from 0.45% (95% CI 0.23-0.67) for individuals with low PRS and low PCE to 14.6% (95% CI 12.8-15.5) for individuals with high PRS and high PCE. The stratification decreased but persisted in the older age groups (**B** and **C**). Here we feature the same Y axis to emphasize differences in *absolute* risk among young, middle and older individuals. **CAD**: coronary artery disease, **PRS**: polygenic risk score, **PCE**: Pooled Cohort Equations.

We then compared the ability of a high PRS vs. high PCE in predicting CAD events across different age groups (Figure 4A). At younger ages of enrollment (40-45 years), high PRS predicted over 3.5-fold more events compared to high PCE – 32.3% (95% CI 32.0-32.5) of CAD events occurring in this age group were predicted by high PRS alone compared to only 9.1% (95% CI 9.0-9.2) by high PCE alone.

**Figure 4.**
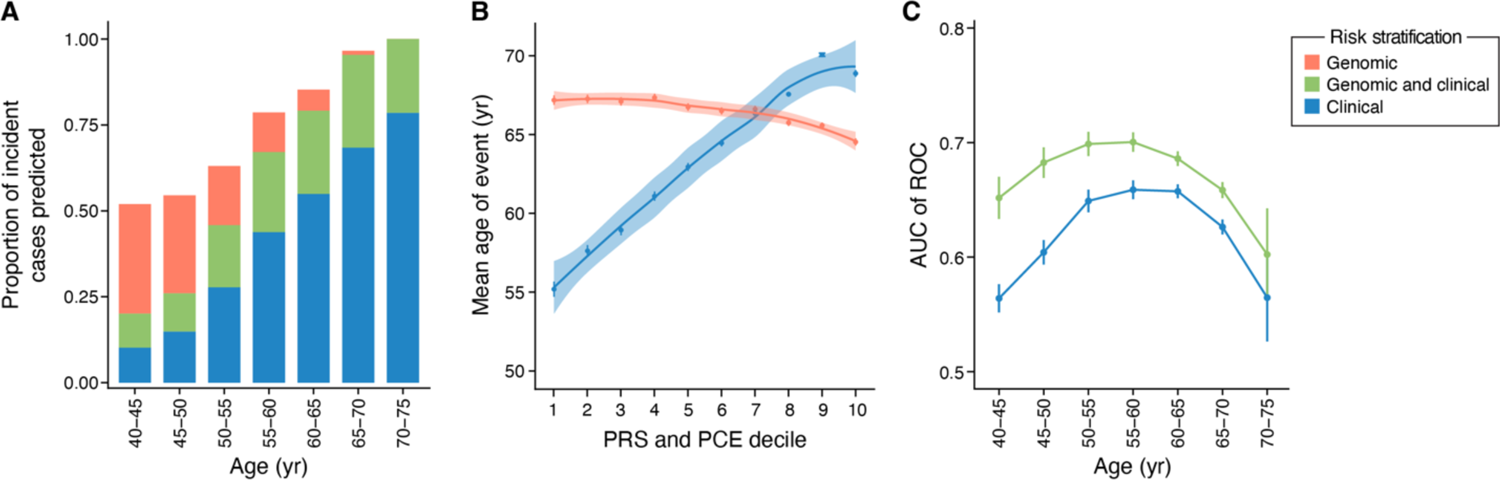
Augmenting Risk Prediction of CAD in Early Middle-Age with the Addition of PRS In the UK Biobank (N=327,837), we show is the proportion of cumulative CAD events predicted using high genomic risk (PRS in the top quintile), intermediate to high clinical risk (PCE 10-year risk ≥7.5%) or both at enrollment, by age of estimation. **B.** Mean age of CAD event decreased with increasing PRS (red), from 67.2 (95% CI 66.6-67.8) years in the lowest decile to 64.5 (95% CI 64.1-65.0) years in the highest decile. Conversely, those in the highest PCE decile (blue) had events 13.7 years later in life than those of the lowest PCE**. C.** AUC of a model considering clinical risk only when compared to a combined clinical and genomic risk model for participants in 5-year age strata between ages 40 and 75 years at age of risk estimation. Genomic risk categories are defined as PRS in the top quintile, middle three quintiles, and bottom quintiles. Clinical risk categories are defined by PCE predicted 10-year risk <7.5%, 7.5-20%, and > 20%. **CAD**: coronary artery disease, **PRS**: polygenic risk score, **PCE**: Pooled Cohort Equations. **AUC**: Area Under the Receiver Operator Curve

### Prediction of premature CAD events

Individuals with high PRS developed CAD earlier in life (mean 65.3 [95% CI 65.0-65.5] years), whereas the average age of first CAD among high PCE group was 70.8 [95% CI 70.6-71.0] years (Supplementary Tables 7 and 8). Mean age of CAD event decreased with increasing PRS, from 67.2 (95% CI 66.6-67.8) years in the lowest decile to 64.5 (95% CI 64.1-65.0) years in the highest decile. Conversely, individuals in the highest PCE decile had events 13.7 years later in life than those of the lowest PCE (Figure 4B, Supplementary Table 9, Supplementary Figure 8). Among individuals with CAD events occurring at less than 55 years, 427 (39.3%) had high PRS but only 32 (2.9%) had high PCE.

### Augmenting clinical risk models with genomic risk

Adding PRS to PCE augmented AUC across all ages but with the greatest impact in younger individuals (Figure 4C, Supplementary Table 10). For individuals <55 years, the improvement was 6.3% (95% CI 4.8-7.8) compared to only 2.9% (95% CI 2.2-3.8) for those over 55.

Furthermore, the AUC increased by 8.8% (95% CI 8.4-9.2%) in the 40-45 age group, 7.8% (95% CI 7.6-8.0%) in the 45–50-year group, and 4.9% (95% CI 4.7-5.1%) in the 50-55 age group, respectively (Figure 4C). The net proportion of CAD cases correctly reclassified by genomic risk (high PRS) was the highest in younger participants (16.1% for age <50 years and 3.4% for age <55 years) but receded for those over 55. The net proportion of controls correctly reclassified by genomic risk (low PRS) was the highest at older ages (15.1% at age <75 years) but diminishes in utility for those younger than 60 (Supplementary Figure 9, Supplementary Table 11).

### Discussion

Our findings enhance our understanding of CAD risk factors by illustrating their dynamic importance throughout life. Unlike traditional models that operate under the constraints of fixed windows of time and proportional hazards, our work goes beyond these limitations to embrace the time-varying nature of these risk factors. The ability to track this dynamic trajectory provides new granularity in risk assessment, particularly for younger individuals. Our approach not only reconciles the time-varying impact of genomic and clinical risk factors but also highlights that CAD PRS offers value for risk assessment in individuals under 55 years over clinical risk factors alone.

While current risk stratification emphasizes a focus on short-term risk, even an emphasis on a longer duration of risk fails to capture the dynamic trajectory of an individual’s changing risk profile over time. Our dynamic model of both genomic and clinical risk factors offers several practical implications. First, it is more accurate than existing risk calculators based on clinical risk factors alone. Second, it allows for more precise clinical risk stratification among younger individuals, for whom clinical risk factors perform least well. Lastly, our work supports the integration of genomics into clinical practice toward improved prevention of premature CAD events, which are generally missed by current clinical risk calculators.

Hazard ratios for conventional CAD risk factors and PRS are both age-dependent and challenges traditional modeling assumptions. This is important for consideration of risk across the life course beyond the present 10-year estimated risk framework, as recently highlighted in a National Heart Lung and Blood Institute workshop.^18^ The Cox proportional hazards model has been the default approach for cardiovascular risk prediction, but its fundamental assumption – that the hazards in both groups compared are proportional – is often erroneous, and commonly reported hazard ratios and risk estimates, such as the ten-year risk estimate from the PCE, are weighted averages of time-varying hazard ratios.^19^

Current risk calculators provide a fixed window estimate, as opposed to a dynamic trajectory.^20,5^ An incremental enhancement to the existing approach would include the use of time-varying covariates and time-varying effects.^21,22^ The Cox-related approach requires the specification of repeated measures of a particular risk factor, which can be challenging to obtain in practice and is often confounded by frequency of ascertainment. The alternative, using time-varying effects,^22^ is an improvement, but the interpretation of these estimates is altogether different: each estimate represents the hazard ratio of a particular covariate on risk within a respective finite time interval, whereas our approach describes the average overall hazard for an individual of a particular age at prediction and thus a particular age period, which is more clinically relevant. We emphasize that genomics allows us to predict lifetime risk early and not only premature events. While the PCE tends to capture individuals who have higher rates of known clinical risk factors, genetic risk is largely independent with a broadly uniform distribution of clinical risk factors among varying levels of genetic risk. PCE incorporates age as a constant interaction with time-to-risk models but our study shows that this change is not linear nor easily predictable.^5,3^ Finally, while the advent of machine learning has opened the possibility of deep-learning for predictive algorithms on much larger data sets, interpretable models that can be feasibly incorporated and understood within the confines of a short clinical visit are essential.^23^ Future approaches need to account for time-varying effects while also considering the time of assessment. This may require the use of time-varying coefficients,^24^ multistate models,^25^ and a more nuanced approach to handling time-varying competing risks.^26^

In conclusion, our work highlights three areas in which CAD PRS adds value to current guideline-based clinical risk prediction using the PCE: (i) CAD PRS had the most value in augmenting risk prediction for CAD among individuals younger than 55 years of age. Prior work for CAD has largely examined AUC augmentation with PRS in aggregate of middle-aged or even older participants noting minimal incremental value.^27,28^ (ii) CAD PRS improves precision in risk estimation for individuals within the strata of clinical risk according to the PCE throughout the life course, but that such stratification is highest among individuals under age 55 years. (iii) Integration of genomics in risk prediction enables the detection of premature events that are missed by current guideline-supported tools. Collectively, these findings support inclusion of PRS to augment current clinical risk estimation toward better allocation of preventive therapies.^29,30^

### Limitations

Our results should be interpreted in the context of potential limitations. First, survival bias is an important limitation with a broad age of inclusion in any volunteer cohort. However, this also reflects the dynamic importance of risk factors when considering event-free individuals at increasing age, which is leveraged in the present study. Second, the two cohorts studied spanned different countries, time periods, and medical guidelines epochs, making absolute estimates between FOS and UKB not directly comparable, but the overall dynamic age-dependent trends were consistent. Third, we do not compare genomic to lifestyle-based “primordial” risk calculators in individuals under the age of 40 year, which would further illuminate the value of genomics in comparison to those measures prior to onset of disease risk factors. Fourth, because this study is predominantly of individuals of European ancestry, additional research is needed to evaluate whether these observations are applicable to other ancestries. CAD PRS has reduced performance in ancestries outside of Europe but cross-ethnic transferability of PRS is improving with more diverse training data and novel methods.^31^

## Conclusions

In summary, this study extends current CAD risk prediction models by offering a dynamic framework that also includes genomics toward improved prediction. We show that genomic information adds the most information for young and middle-aged individuals when compared with older individuals for the prediction of CAD events.

## Data Availability

UK Biobank data is available through application to the UK Biobank. Framingham Offspring Cohort Data is available through application to dbGAP. All code to reproduce analysis is available at surbut.github.io/dynamichr.

## Acknowledgments

We would like to thank the participants of the FOS and UKB. Additionally, we would like to thank Dr. Francesca Domenici, Dr. Giovanni Parmigiani, Dr. Sasha Gusev and Dr. Ludovic Trinquart for the valuable input on statistical approaches for lifetime risk modeling. We would like to thank Leslie Gaffney at the Broad Communications Team for her invaluable support and counsel on figure revision.

## Non-standard Abbreviations and Acronyms

CAD: Coronary artery disease

FOS: Framingham Offspring Study

UKB: UK Biobank

PRS: Polygenic risk score

PCE: Pooled Cohort Equations

LDL-C: Low-density lipoprotein cholesterol

ASCVD: Atherosclerotic cardiovascular disease

