## Supplemental Figures and Tables for "Dynamic Importance of Genomic and Clinical Risk for Coronary Artery Disease Over the Life Course"

Urbut SM et al.

**Outline**

**Supplementary Figures**

Supplementary Figure 1: Selection of study participants from the Framingham Offspring Study

Supplementary Figure 2: Selection of study participants from the UK Biobank

Supplementary Figure 3: Non-proportional hazards of risk factors for CAD

Supplementary Figure 4: Hazard ratio and proportion of variation explained by Pooled Cohort Equations nominal 10-year ASCVD risk percentile and standard deviation in the UK Biobank (N=327,837)

Supplementary Figure 5: Distribution of predicted 10-year risk percentile in the UK Biobank by age (N=327,837)

Supplementary Figure 6: Proportion of variation explained by different genomic and clinical CAD risk factors by age at estimation in the Framingham Offspring Cohort and the UK Biobank

Supplementary Figure 7: Distribution of age by genomic risk category in the UK Biobank (N=327,837)

Supplementary Figure 8: Cumulative hazard of CAD over the life course in the UK Biobank (N=327,837)

Supplementary Figure 9: Net reclassification of CAD events and non-events with genomic risk at different age cut-offs in the UK Biobank (N=327,837)

**Supplementary Tables**

Supplementary Table 1: Age-dependent hazard ratio for genomic and nongenomic risk factors in the FOS (N=3,558)

Supplementary Table 2: Age-dependent hazard ratio for genomic and nongenomic risk factors in the UKB (N=327,837)

Supplementary Table 3: Age-dependent proportion of variation explained by genomic and nongenomic risk factors in the FOS (N=3,660)

Supplementary Table 4: Age-dependent proportion of variation explained by genomic and nongenomic risk factors in the UKB (N=327,837)

Supplementary Table 5: Translation of age-specific PCE clinical risk quintiles to predicted 10-year ASCVD risk (N=327,837)

Supplementary Table 6: Proportion of CAD events predicted by genomic and nongenomic risk by age of estimation in the UKB (N=327,837)

Supplementary Table 7: Characteristics of study participants by genomic risk category in the UKB (N=327,837)

Supplementary Table 8: Characteristics of study participants by clinical risk category in the UKB (N=327,837)

Supplementary Table 9: Mean age of CAD event by genomic or clinical risk decile in the UKB (N=327,837)

Supplementary Table 10: Discrimination of clinical vs. combined clinical-genomic risk models by age in the UKB (N=327,837)

Supplementary Table 11: Net reclassification of CAD events and non-events with the CAD PRS by age in the UK Biobank (N=327,837)

Supplementary Table 12: Proportion of individuals by age in each predicted 10-year ASCVD risk category in the UKB (327,837).

**Supplementary Methods**

**Supplementary Figure 1: Selection of study participants from the Framingham Offspring Study**

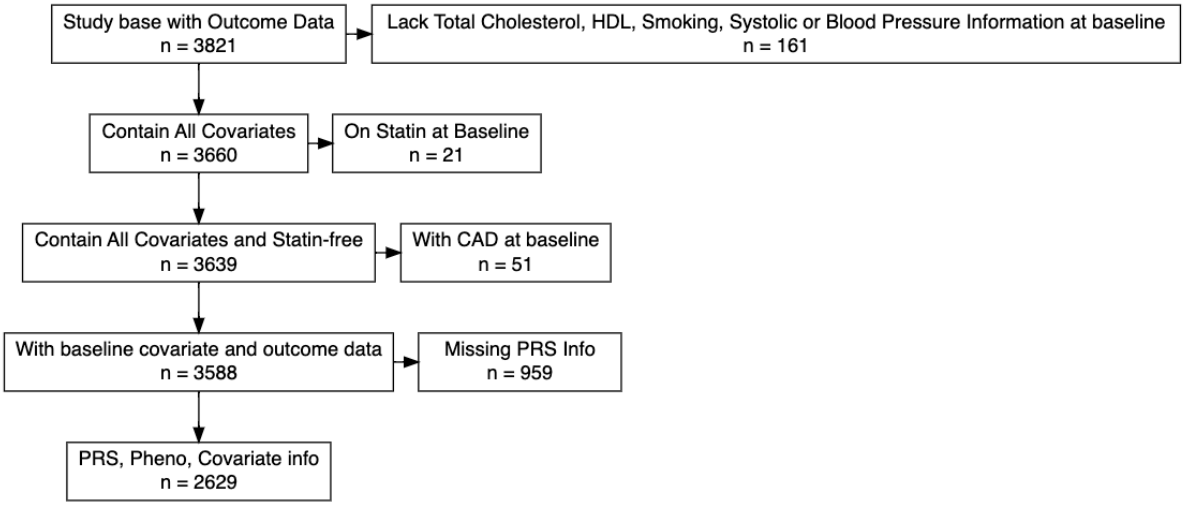

Out of a total of 3,821 individuals form the Framingham Offspring Study (FOS), we excluded 161 participants with missing risk factor (covariates) data necessary for analysis, 21 on lipid-lowering medication at baseline, and 51 with prevalent coronary artery disease at baseline. The 3,588 individuals remaining were used for analysis of clinical risk factors. From this group, there were 959 participants who did not have genomic information, keeping a subset of 2,628 individuals used for the genetic analysis. CAD: coronary artery disease, PRS: polygenic risk score, HDL: high-density lipoprotein cholesterol

**Supplementary Figure 2: Selection of study participants from the UK Biobank**

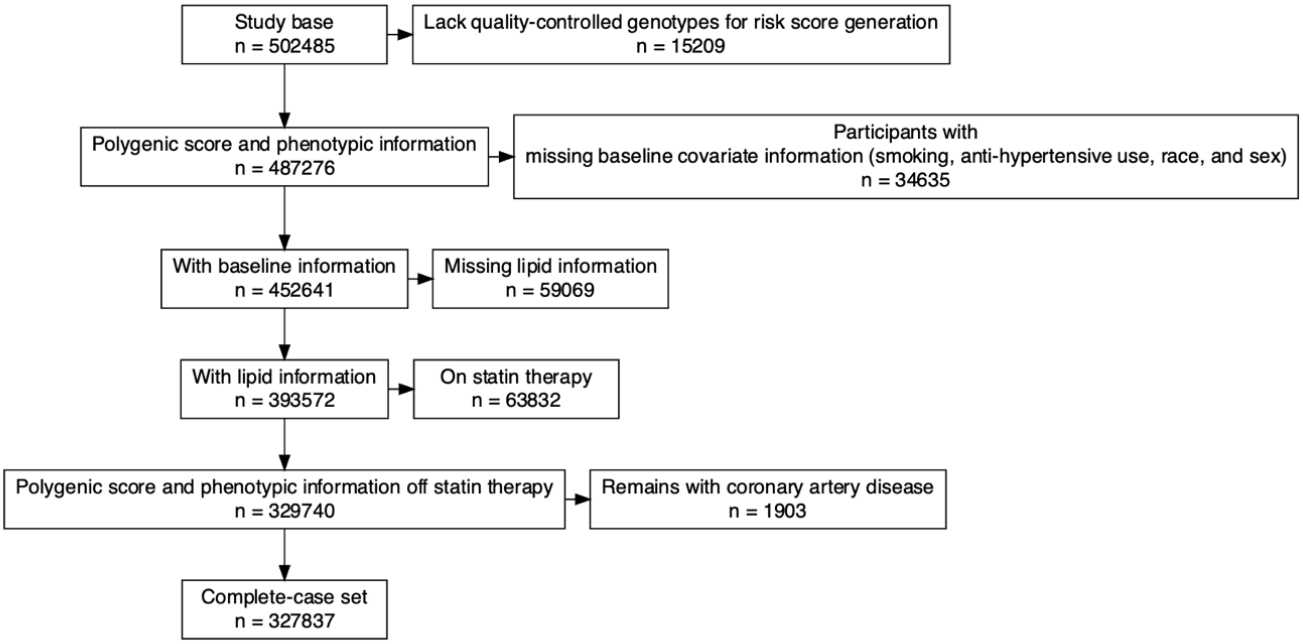

Out of total 502,485 participants in the UK Biobank (UKB), we excluded 15,209 who lacked quality-controlled genomic information necessary for polygenic risk score calculation, 34,635 participants with missing covariate information for clinical risk calculation, 49,069 with missing lipid levels, 63,832 on statin therapy at baseline, and 1,903 remaining with coronary artery disease at baseline. The final dataset used for analysis after those exclusions was 327,837 participants.

**Supplementary Figure 3: Non-proportional hazards of risk factors for CAD**

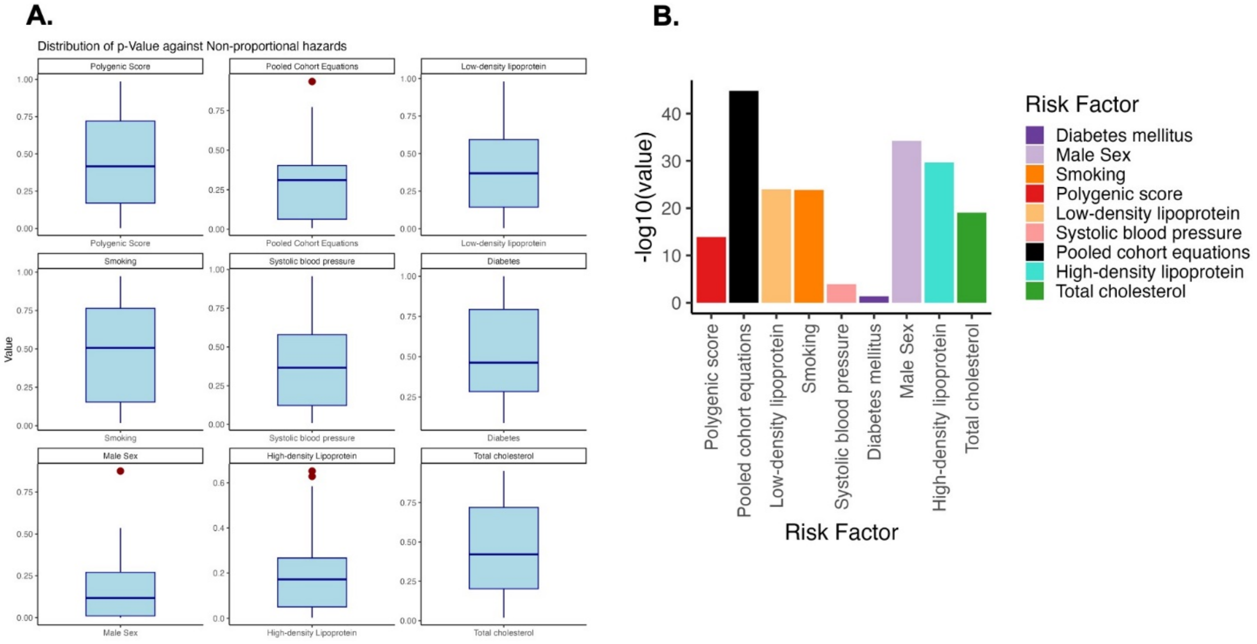

In the UKB (N=327,837) we test the Schoenfeld residuals assumption in which the global null posits that the trend in residuals over time is not-significantly different than zero. Under our model (A) in which the stratification occurs for each age of enrollment, here we display the distribution of p-values over all ages considered between 41 and 70 years. Each boxplot displays the distribution of p values for sequential fits of individuals at each age of enrollment. In (B) and under a model which does not account for change in risk factor importance over time, we assessed the fit of a typical Cox proportional hazards without stratification by age by assessing Schoenfeld residuals over the time period considered. In summary we demonstrate that the cox proportional hazards assumption is violated for all covariates considered and resolved by a model that stratifies by time of estimation as compared to one that stratifies by time of event, and more clinically-actionable for a provider meeting a patient of a given age.

**Supplementary Figure 4: Hazard ratio and proportion of variation explained by Pooled Cohort Equations nominal 10-year risk percentile and standard deviation in the UK Biobank (N=327,837)**

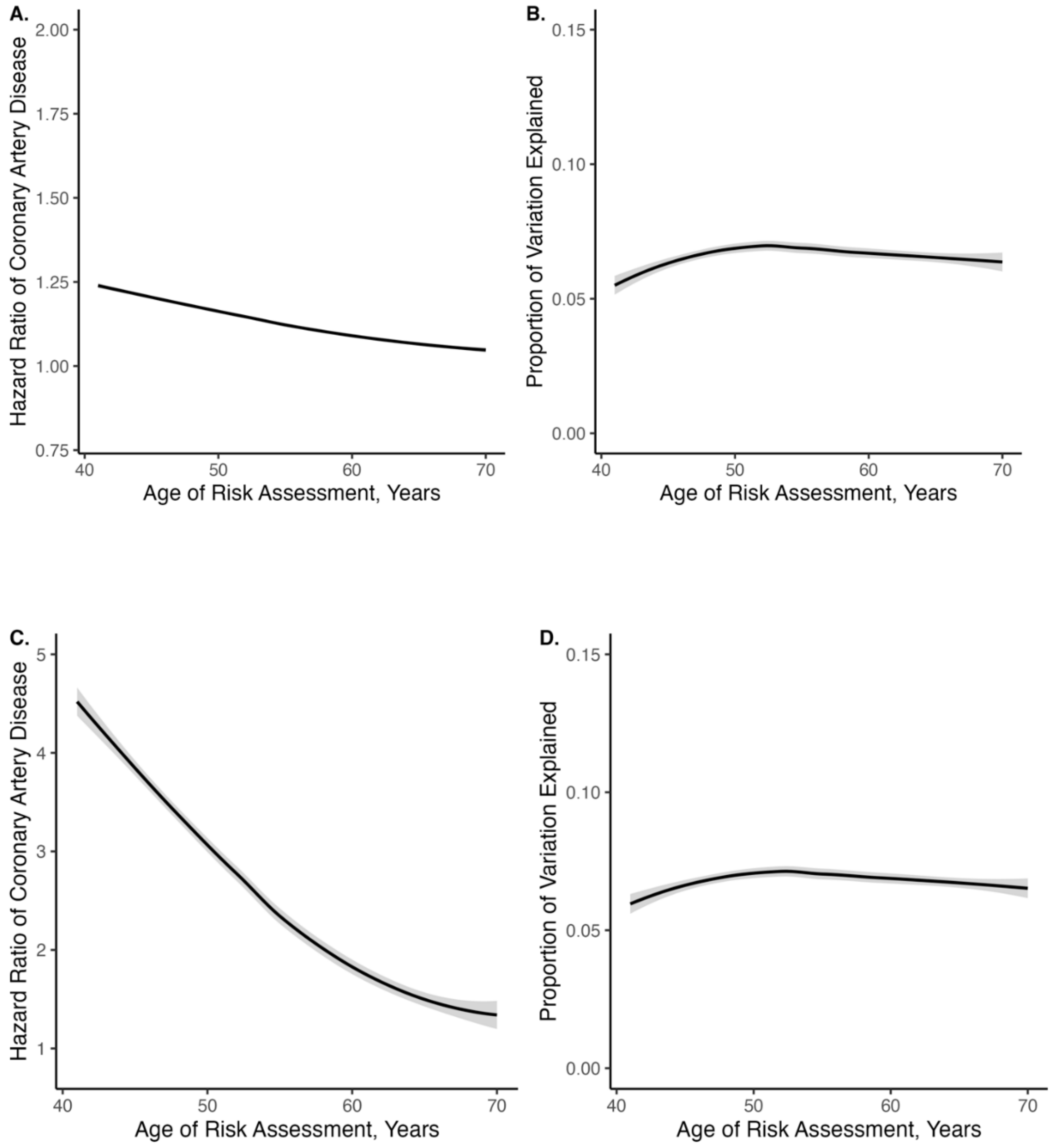

We display the age-dependent hazard ratio using the overall predicted PCE 10-year ASCVD risk percentile (A,B) and PCE scaled by their overall standard deviation (one standard deviation is equivalent to 7.2% 10-year ASCVD risk) (C,D) PCE: Pooled Cohort Equations, ASCVD: Atherosclerotic Cardiovascular Disease

**Supplementary Figure 5: Distribution of predicted 10-year risk percentile in the UK Biobank by age (N=327,837)**

**A.**

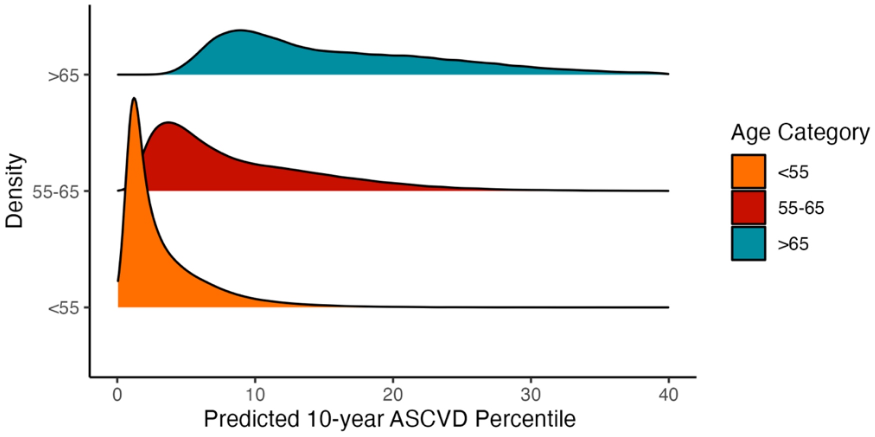

**B.**

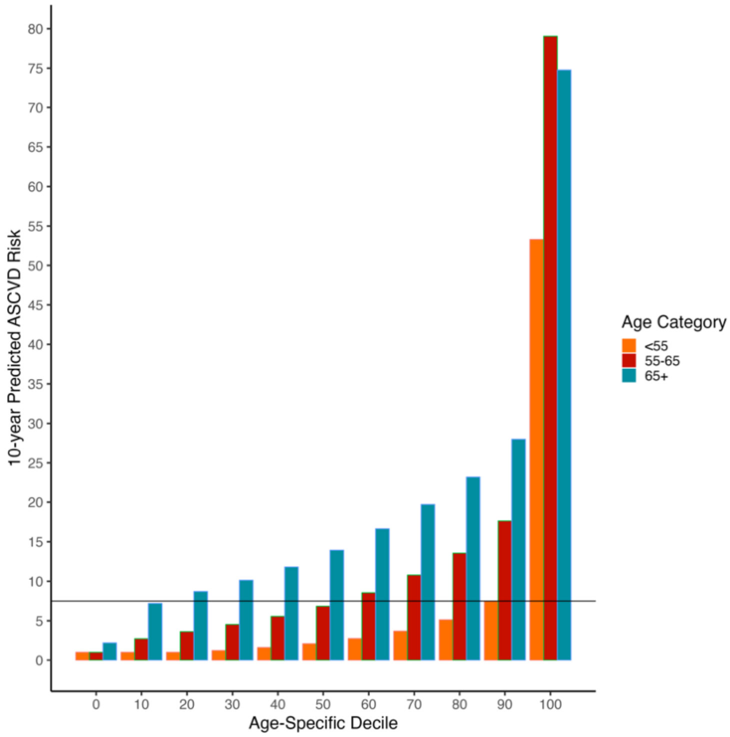

In the UKB (N=327,837), we show the distribution of predicted 10-year ASCVD risk by PCE for each age category (A) and translation of age-specific decile to predicted 10-year ASCVD risk (B). Horizontal line indicates 7.5% predicted 10-year ASCVD risk. PCE: Pooled Cohort Equations, ASCVD: Atherosclerotic Cardiovascular Disease

**Supplementary Figure 6: Distribution of age by genomic risk in the UK Biobank (N=327,837)**

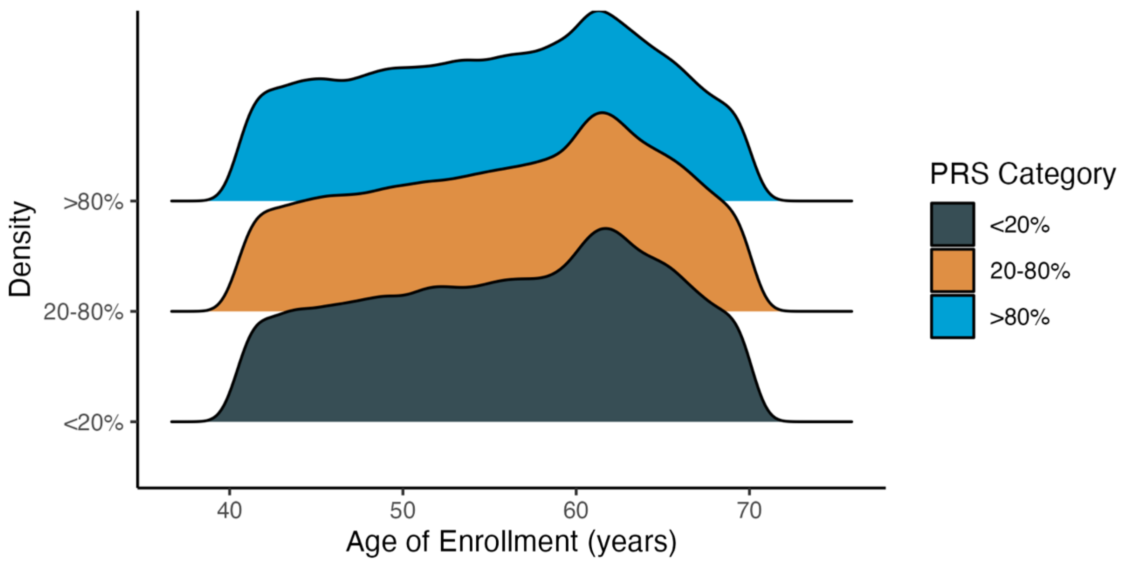

In the UKB (N=327,837) we display the distribution of enrollment ages by genomic risk category for low (<20%), intermediate (20-80%) and high (>80%) genomic risk to demonstrate that no genomic risk class is disproportionately enriched in one age distribution in this primary prevention cohort of the UKB. **PRS:** Polygenic Risk Score.**UKB:** UK Biobank.

**Supplementary Figure 7:** Proportion of variation explained by different genomic and clinical CAD risk factors by age at estimation in the Framingham Offspring Cohort and the UK Biobank (N=327,827)

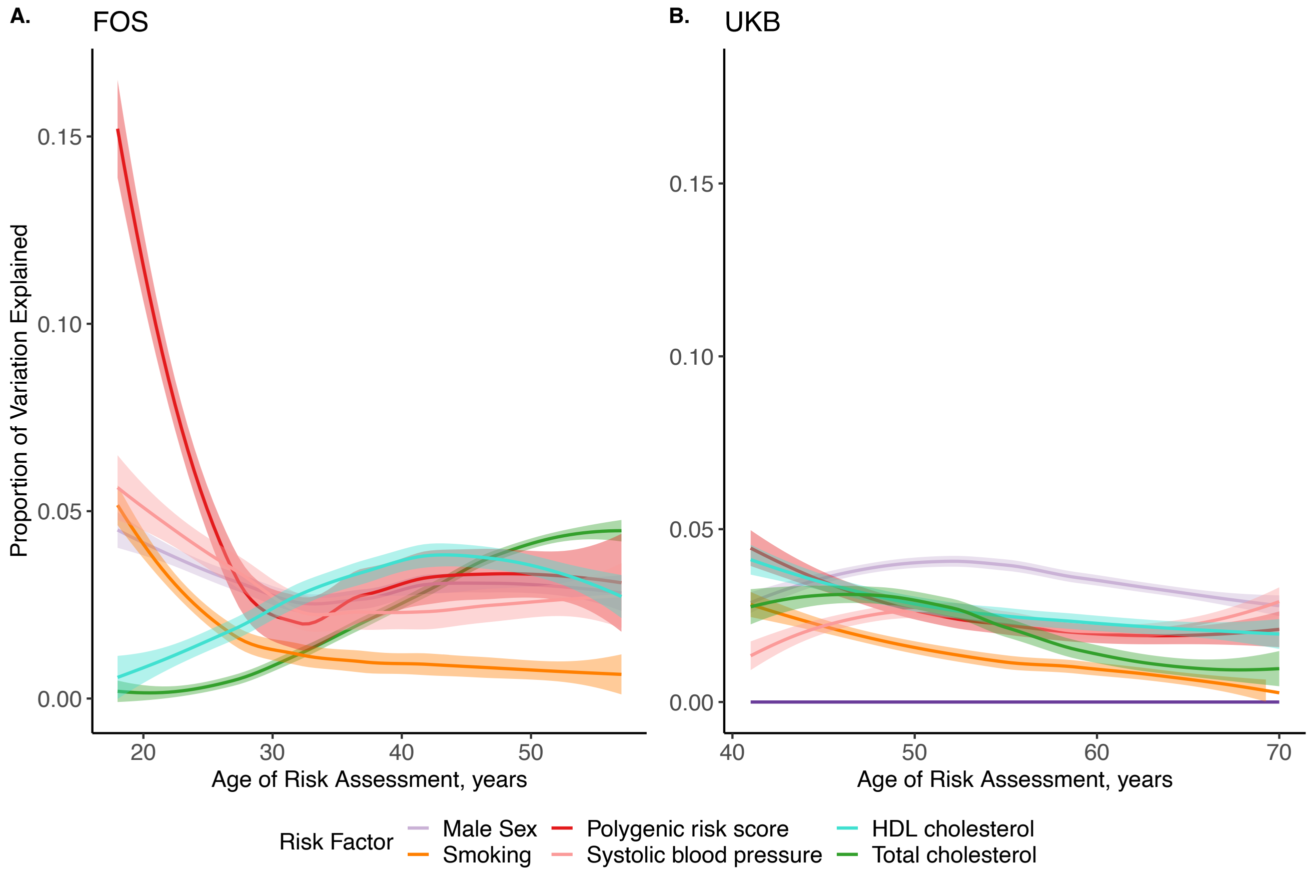

The proportion of variation explained (PVE) by multiple risk factors is plotted at each age of enrollment (A) between 18 and 57 years in the Framingham Offspring Cohort (FOS) and (B) between 41 and 70 years in the UK Biobank. The PVE was obtained from McFadden's Pseudo R2 from a logistic regression model estimating the effect of each risk factor on the development of coronary artery disease for individuals up to and including the age of risk assessment. No additional covariates are used in the analysis to isolate the effect of each risk factor separately. The PVE by diabetes is not shown in FOS due to low incidence and insufficient counts to generate estimates. The PVE by diabetes in the UKB appears flat due to very low proportions of individuals in the population (all values are <0.2% and are shown in Supplementary Table 4). **PVE**: proportion of variation explained (PVE. **FOS:** Framingham Offspring Cohort, **UKB**: UK Biobank.

**Supplementary Figure 8: Cumulative hazard of CAD over the life course in the UK Biobank (N=327,837)**

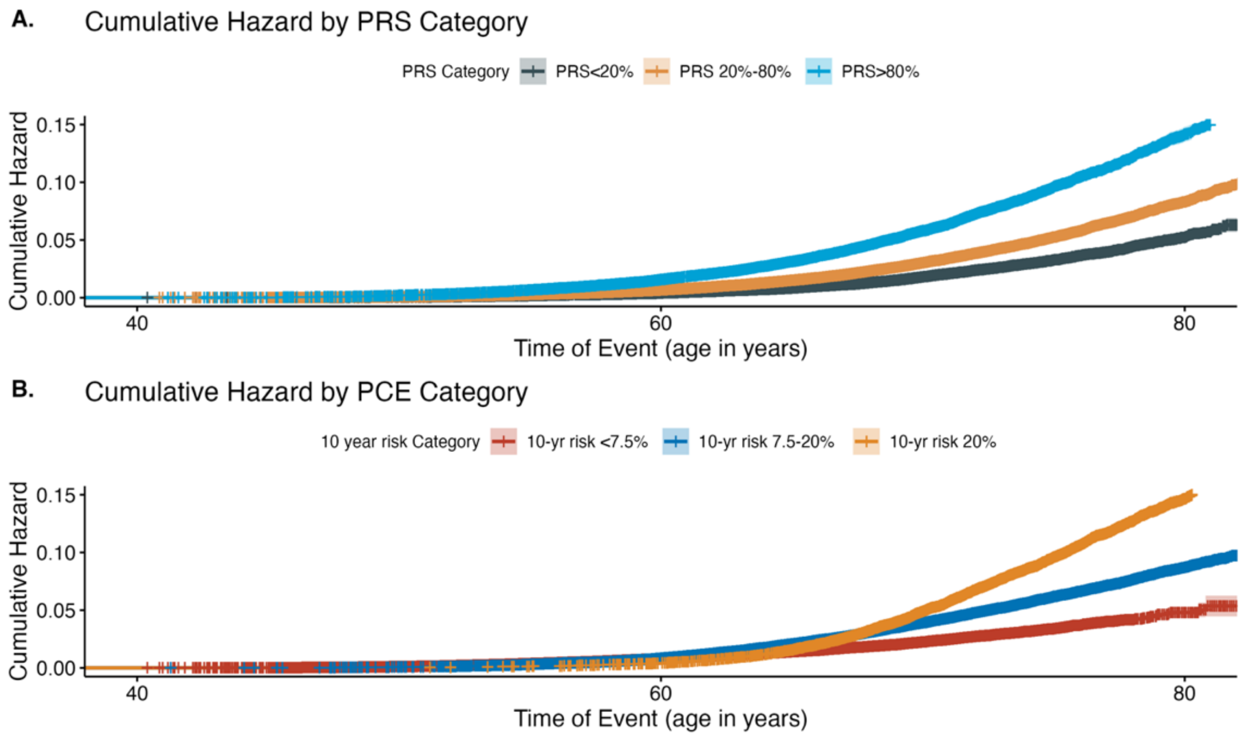

In the UK Biobank (N=327,837) we show the cumulative hazard of CAD diagnosis by age of event (A) for genomic risk categories – low (PRS in bottom quintile), intermediate (PRS middle three quintiles), and high (PRS in top quintile) and (B) for clinical risk categories – PCE 10-year ASCVD risk <7.5%, between 7.5% and 20%, and >20%. CAD events are stratified earlier in the life course by genomic risk compared to clinical risk. There are two key advantages of using age scale. First, within the model all comparisons involve pairs of individuals that are the same age, allowing for non-parametric estimates of the importance of each variable with age. Second, the description extent is more than doubled from age 40 to 75 (i.e. 35 years) as opposed to the maximum time from enrollment to outcome events (i.e. 12.1 years). **PRS:** Polygenic Risk Score, **PCE:** Pooled Cohort Equations, **ASCVD:** Atherosclerotic Cardiovascular Disease

**Supplementary Figure 9: Net reclassification of CAD events and non-events with genomic risk at different age cut-offs in the UKB (N=327,837)** .

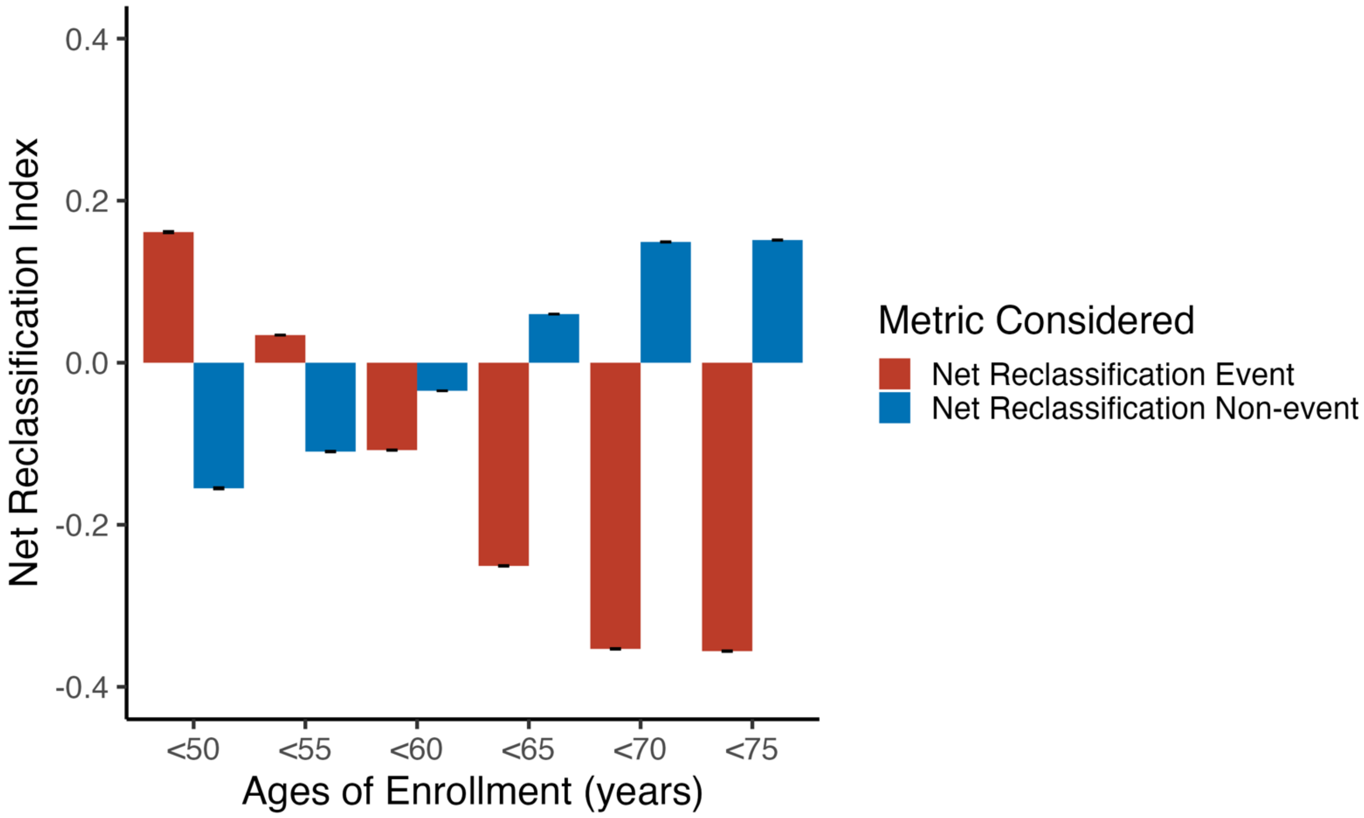

Here, we compute the net number of events (red) correctly reclassified using a high genomic risk (PRS in the top quintile) versus clinical risk (PCE 10-year ASCVD risk >20%) in red. We also report the net non-events (blue) reclassified using a low genomic risk (PRS in the bottom quintile) when compared to low clinical risk (PCE 10-year ASCVD risk <7.5%).
**PRS:** Polygenic Risk Score, **PCE**: Pooled Cohort Equations, **ASCVD**: Atherosclerotic Cardiovascular Disease, **NRI:** Net reclassification index.

**Supplementary Table 1**: Age-dependent hazard ratio for genomic and nongenomic risk factors in the FOS between ages (N=3,588)

| Age (years) | PRS, HR (95% CI) | Smoking, HR (95% CI) | SBP, HR (95% CI) | TC, HR (95% CI) | HDL-C, HR (95% CI) | Male Sex, HR (95% CI) |
|---|---|---|---|---|---|---|
| 18.00 | 2.68 (0.99-7.25) | 3.15 (0.53-18.86) | 1.81 (0.81-4.06) | 0.43 (0.09-2.03) | 0.45 (0.13-1.58) | 4.65 (0.52-41.65) |
| 19.00 | 3.58 (1.39-9.19) | 1.98 (0.44-8.84) | 1.95 (0.96-3.95) | 0.45 (0.14-1.44) | 0.47 (0.16-1.40) | 3.29 (0.64-16.95) |
| 20.00 | 1.77 (0.72-4.32) | 1.50 (0.34-6.71) | 2.18 (1.05-4.56) | 0.83 (0.30-2.29) | 0.93 (0.37-2.34) | 2.78 (0.54-14.35) |
| 21.00 | 1.65 (0.70-3.85) | 1.40 (0.31-6.25) | 2.85 (1.09-7.47) | 1.10 (0.41-2.99) | 1.52 (0.66-3.48) | 2.79 (0.54-14.38) |
| 22.00 | 1.38 (0.68-2.78) | 1.52 (0.43-5.38) | 2.68 (1.12-6.42) | 1.14 (0.57-2.29) | 0.87 (0.40-1.90) | 4.82 (1.02-22.72) |
| 23.00 | 1.62 (0.81-3.23) | 2.85 (0.76-10.73) | 2.44 (1.18-5.05) | 0.70 (0.31-1.60) | 0.66 (0.30-1.49) | 4.20 (1.11-15.82) |
| 24.00 | 1.58 (0.99-2.51) | 1.79 (0.76-4.23) | 1.57 (0.95-2.58) | 0.99 (0.61-1.60) | 0.44 (0.26-0.74) | 3.29 (1.41-7.69) |
| 25.00 | 1.20 (0.80-1.80) | 1.59 (0.80-3.14) | 1.26 (0.82-1.94) | 1.03 (0.71-1.50) | 0.69 (0.46-1.02) | 2.78 (1.36-5.67) |
| 26.00 | 1.15 (0.81-1.64) | 1.78 (0.92-3.44) | 1.30 (0.88-1.92) | 1.16 (0.83-1.61) | 0.64 (0.44-0.92) | 2.97 (1.48-5.97) |
| 27.00 | 1.53 (1.15-2.03) | 1.91 (1.10-3.32) | 1.48 (1.07-2.05) | 1.15 (0.86-1.54) | 0.62 (0.45-0.84) | 2.14 (1.22-3.75) |
| 28.00 | 1.43 (1.10-1.86) | 2.07 (1.18-3.64) | 1.50 (1.10-2.05) | 1.29 (0.95-1.77) | 0.60 (0.43-0.82) | 1.92 (1.11-3.33) |
| 29.00 | 1.52 (1.16-1.99) | 1.93 (1.15-3.26) | 1.40 (1.03-1.91) | 1.45 (1.08-1.95) | 0.56 (0.41-0.76) | 1.94 (1.15-3.28) |
| 30.00 | 1.28 (0.96-1.70) | 1.51 (0.90-2.53) | 1.57 (1.16-2.12) | 1.67 (1.29-2.15) | 0.51 (0.37-0.71) | 2.39 (1.39-4.12) |
| 31.00 | 1.64 (1.23-2.19) | 1.44 (0.88-2.36) | 1.79 (1.32-2.42) | 1.53 (1.19-1.95) | 0.54 (0.40-0.74) | 2.36 (1.40-3.96) |
| 32.00 | 1.50 (1.13-2.00) | 1.19 (0.73-1.93) | 1.61 (1.27-2.05) | 1.50 (1.17-1.93) | 0.61 (0.46-0.82) | 2.02 (1.23-3.32) |
| 33.00 | 1.56 (1.15-2.13) | 1.48 (0.89-2.46) | 1.45 (1.11-1.89) | 1.36 (1.02-1.81) | 0.71 (0.54-0.94) | 2.35 (1.38-4.00) |
| 34.00 | 1.64 (1.22-2.19) | 1.55 (0.95-2.51) | 1.36 (1.05-1.75) | 1.46 (1.11-1.92) | 0.62 (0.47-0.82) | 1.66 (1.02-2.70) |
| 35.00 | 1.90 (1.44-2.50) | 2.46 (1.53-3.96) | 1.27 (0.96-1.68) | 1.41 (1.10-1.79) | 0.50 (0.38-0.66) | 2.56 (1.56-4.21) |
| 36.00 | 1.96 (1.47-2.63) | 2.29 (1.41-3.73) | 1.34 (1.01-1.78) | 1.52 (1.17-1.98) | 0.55 (0.41-0.74) | 1.92 (1.17-3.16) |
| 37.00 | 1.73 (1.30-2.30) | 2.11 (1.32-3.39) | 1.38 (1.09-1.75) | 1.69 (1.32-2.17) | 0.54 (0.41-0.72) | 3.91 (2.20-6.92) |
| 38.00 | 1.60 (1.20-2.13) | 1.83 (1.14-2.95) | 1.47 (1.17-1.85) | 1.91 (1.48-2.47) | 0.54 (0.40-0.72) | 2.92 (1.70-5.00) |
| 39.00 | 1.51 (1.12-2.02) | 1.74 (1.09-2.76) | 1.42 (1.14-1.77) | 1.55 (1.20-2.02) | 0.57 (0.44-0.75) | 3.60 (2.09-6.22) |
| 40.00 | 1.58 (1.20-2.09) | 1.98 (1.26-3.09) | 1.38 (1.09-1.75) | 1.23 (0.95-1.58) | 0.51 (0.38-0.67) | 3.25 (2.00-5.26) |
| 41.00 | 1.62 (1.25-2.11) | 1.71 (1.12-2.60) | 1.47 (1.19-1.83) | 1.31 (1.03-1.68) | 0.52 (0.40-0.68) | 2.94 (1.87-4.62) |
| 42.00 | 1.60 (1.24-2.07) | 1.78 (1.19-2.67) | 1.43 (1.16-1.77) | 1.48 (1.20-1.82) | 0.41 (0.31-0.53) | 2.84 (1.85-4.37) |
| 43.00 | 1.60 (1.20-2.13) | 1.33 (0.85-2.08) | 1.34 (1.07-1.70) | 1.70 (1.39-2.08) | 0.49 (0.38-0.63) | 2.85 (1.78-4.55) |
| 44.00 | 1.83 (1.37-2.43) | 1.78 (1.16-2.73) | 1.14 (0.91-1.42) | 1.45 (1.17-1.78) | 0.50 (0.39-0.64) | 2.62 (1.69-4.07) |
| 45.00 | 1.86 (1.42-2.42) | 1.70 (1.13-2.56) | 1.15 (0.93-1.41) | 1.59 (1.30-1.95) | 0.63 (0.51-0.79) | 2.04 (1.35-3.08) |
| 46.00 | 1.66 (1.27-2.17) | 2.34 (1.56-3.53) | 1.24 (1.03-1.48) | 1.63 (1.33-1.99) | 0.71 (0.57-0.88) | 1.58 (1.06-2.36) |
| 47.00 | 1.36 (1.05-1.78) | 2.00 (1.34-2.98) | 1.32 (1.10-1.57) | 1.70 (1.42-2.04) | 0.80 (0.65-0.99) | 1.85 (1.24-2.77) |
| 48.00 | 1.34 (1.00-1.79) | 1.93 (1.30-2.88) | 1.34 (1.13-1.60) | 1.57 (1.30-1.89) | 0.67 (0.54-0.84) | 2.42 (1.58-3.71) |
| 49.00 | 1.53 (1.18-1.98) | 1.52 (1.03-2.25) | 1.25 (1.05-1.48) | 1.39 (1.16-1.67) | 0.63 (0.50-0.79) | 2.03 (1.35-3.05) |
| 50.00 | 1.77 (1.33-2.37) | 1.36 (0.90-2.07) | 1.23 (1.03-1.47) | 1.17 (0.96-1.42) | 0.72 (0.58-0.89) | 1.91 (1.24-2.95) |
| 51.00 | 1.58 (1.15-2.17) | 1.46 (0.93-2.28) | 1.23 (1.03-1.47) | 1.13 (0.92-1.39) | 0.79 (0.63-0.99) | 1.49 (0.94-2.37) |
| 52.00 | 1.80 (1.19-2.72) | 1.42 (0.86-2.34) | 1.11 (0.92-1.33) | 1.06 (0.83-1.34) | 0.76 (0.60-0.98) | 2.94 (1.59-5.44) |
| 53.00 | 1.32 (0.79-2.20) | 1.32 (0.76-2.32) | 1.02 (0.83-1.24) | 1.10 (0.85-1.42) | 0.66 (0.50-0.88) | 2.35 (1.25-4.43) |
| 54.00 | 2.00 (1.24-3.22) | 1.10 (0.58-2.10) | 0.94 (0.73-1.19) | 0.89 (0.64-1.23) | 0.65 (0.47-0.88) | 2.61 (1.27-5.37) |
| 55.00 | 1.49 (0.91-2.44) | 1.12 (0.53-2.35) | 1.00 (0.79-1.26) | 0.81 (0.55-1.18) | 0.47 (0.30-0.74) | 1.69 (0.79-3.59) |
| 56.00 | 1.99 (1.07-3.70) | 0.98 (0.41-2.33) | 1.13 (0.86-1.50) | 1.02 (0.67-1.57) | 0.36 (0.20-0.65) | 1.97 (0.80-4.90) |
| 57.00 | 0.88 (0.25-3.16) | 0.90 (0.36-2.25) | 1.16 (0.86-1.58) | 1.18 (0.76-1.84) | 0.26 (0.14-0.51) | 2.59 (0.92-7.25) |

In the Framingham Offspring Study (FOS), we display the age-specific hazard ratio (HR) and 95% confidence interval (CI) per standardized increase in quantitative risk factors, or for binary risk factors, the presence or absence of each.

FOS: Framingham Offspring Study; TC: Total Cholesterol SBP; Systolic Blood pressure; HDL: High density lipoprotein cholesterol, PRS: Polygenic Risk Score

### Supplementary Table 2: Age-dependent hazard ratio for genomic and nongenomic risk factors in the UKB (N=327,837)

| Age (years) | PRS, HR (95% CI) | PCE, HR (95%CI) | Smoking, HR (95% CI) | SBP, HR (95% CI) | DM, HR (95% CI) | Male Sex, HR (95% CI) | HDL-C, HR (95% CI) | TC, HR (95% CI) |
|---|---|---|---|---|---|---|---|---|
| 41 | 2.25 (1.77-2.87) | 1.24 (1.18-1.30) | 3.51 (2.13-5.80) | 1.49 (1.12-1.99) | NA (NA-NA) | 3.20 (1.82-5.64) | 0.38 (0.26-0.54) | 1.68 (1.34-2.11) |
| 42 | 1.73 (1.52-1.96) | 1.22 (1.19-1.25) | 2.82 (2.15-3.69) | 1.69 (1.47-1.94) | 2.40 (0.60-9.67) | 3.71 (2.75-5.00) | 0.43 (0.36-0.51) | 1.75 (1.57-1.96) |
| 43 | 1.64 (1.47-1.84) | 1.22 (1.20-1.25) | 3.11 (2.46-3.94) | 1.71 (1.52-1.92) | 3.16 (1.01-9.86) | 3.88 (2.97-5.06) | 0.45 (0.38-0.53) | 1.79 (1.63-1.97) |
| 44 | 1.58 (1.43-1.75) | 1.21 (1.19-1.23) | 2.98 (2.39-3.72) | 1.79 (1.62-1.99) | 2.36 (0.76-7.36) | 4.11 (3.21-5.25) | 0.47 (0.41-0.54) | 1.86 (1.71-2.03) |
| 45 | 1.68 (1.53-1.84) | 1.21 (1.19-1.23) | 3.09 (2.53-3.78) | 1.82 (1.66-2.00) | 2.72 (1.01-7.27) | 4.30 (3.43-5.40) | 0.48 (0.43-0.55) | 1.79 (1.64-1.94) |
| 46 | 1.60 (1.46-1.75) | 1.20 (1.19-1.22) | 2.81 (2.31-3.41) | 1.77 (1.62-1.94) | 2.09 (0.78-5.59) | 4.01 (3.25-4.94) | 0.53 (0.47-0.59) | 1.70 (1.56-1.84) |
| 47 | 1.66 (1.53-1.81) | 1.20 (1.18-1.22) | 2.64 (2.19-3.19) | 1.69 (1.55-1.84) | 2.78 (1.24-6.21) | 3.37 (2.79-4.08) | 0.52 (0.47-0.58) | 1.61 (1.49-1.74) |
| 48 | 1.67 (1.54-1.82) | 1.20 (1.18-1.21) | 2.43 (2.01-2.93) | 1.65 (1.53-1.79) | 2.65 (1.26-5.59) | 3.26 (2.71-3.92) | 0.51 (0.46-0.56) | 1.61 (1.50-1.74) |
| 49 | 1.67 (1.54-1.81) | 1.18 (1.16-1.20) | 2.49 (2.07-2.99) | 1.63 (1.51-1.76) | 3.17 (1.69-5.92) | 3.29 (2.75-3.92) | 0.52 (0.47-0.57) | 1.56 (1.45-1.68) |
| 50 | 1.66 (1.54-1.79) | 1.16 (1.15-1.18) | 2.44 (2.04-2.92) | 1.63 (1.51-1.76) | 3.09 (1.70-5.61) | 3.55 (2.99-4.23) | 0.56 (0.51-0.61) | 1.52 (1.41-1.63) |
| 51 | 1.61 (1.50-1.73) | 1.15 (1.14-1.16) | 2.41 (2.03-2.86) | 1.52 (1.41-1.63) | 2.41 (1.29-4.50) | 3.67 (3.12-4.33) | 0.58 (0.53-0.63) | 1.42 (1.32-1.52) |
| 52 | 1.54 (1.44-1.65) | 1.14 (1.12-1.15) | 2.20 (1.86-2.60) | 1.48 (1.38-1.58) | 2.32 (1.28-4.21) | 3.80 (3.25-4.44) | 0.57 (0.53-0.62) | 1.39 (1.30-1.49) |
| 53 | 1.50 (1.40-1.60) | 1.13 (1.12-1.14) | 2.43 (2.07-2.84) | 1.42 (1.33-1.52) | 2.00 (1.11-3.63) | 3.63 (3.14-4.21) | 0.60 (0.55-0.65) | 1.33 (1.25-1.42) |
| 54 | 1.56 (1.46-1.66) | 1.13 (1.12-1.14) | 2.43 (2.08-2.84) | 1.42 (1.33-1.51) | 2.83 (1.73-4.65) | 3.57 (3.09-4.12) | 0.58 (0.54-0.63) | 1.28 (1.21-1.37) |
| 55 | 1.62 (1.52-1.73) | 1.12 (1.11-1.13) | 2.44 (2.10-2.83) | 1.42 (1.34-1.51) | 2.66 (1.62-4.36) | 3.34 (2.91-3.83) | 0.58 (0.54-0.62) | 1.25 (1.18-1.33) |
| 56 | 1.58 (1.48-1.68) | 1.12 (1.11-1.12) | 2.30 (1.98-2.68) | 1.43 (1.35-1.52) | 3.10 (1.99-4.82) | 3.38 (2.96-3.86) | 0.54 (0.50-0.58) | 1.21 (1.14-1.28) |
| 57 | 1.54 (1.45-1.63) | 1.11 (1.10-1.11) | 2.31 (2.00-2.66) | 1.44 (1.36-1.52) | 2.78 (1.79-4.32) | 3.22 (2.84-3.65) | 0.55 (0.52-0.59) | 1.18 (1.12-1.25) |
| 58 | 1.55 (1.46-1.64) | 1.11 (1.10-1.11) | 2.27 (1.96-2.61) | 1.40 (1.32-1.47) | 2.77 (1.80-4.26) | 3.34 (2.96-3.76) | 0.53 (0.50-0.57) | 1.11 (1.05-1.17) |
| 59 | 1.56 (1.48-1.65) | 1.10 (1.09-1.11) | 2.07 (1.80-2.38) | 1.40 (1.34-1.47) | 2.50 (1.64-3.81) | 3.03 (2.71-3.38) | 0.54 (0.51-0.58) | 1.12 (1.06-1.18) |
| 60 | 1.45 (1.38-1.52) | 1.09 (1.09-1.10) | 2.12 (1.87-2.41) | 1.39 (1.32-1.45) | 2.90 (2.04-4.12) | 2.96 (2.67-3.28) | 0.55 (0.52-0.58) | 1.09 (1.04-1.15) |
| 61 | 1.40 (1.33-1.46) | 1.09 (1.08-1.09) | 2.07 (1.82-2.34) | 1.36 (1.31-1.42) | 2.88 (2.11-3.94) | 2.81 (2.56-3.09) | 0.59 (0.56-0.62) | 1.10 (1.05-1.15) |
| 62 | 1.38 (1.32-1.45) | 1.08 (1.08-1.09) | 2.04 (1.80-2.31) | 1.34 (1.29-1.40) | 2.66 (1.97-3.58) | 2.86 (2.60-3.13) | 0.61 (0.58-0.64) | 1.06 (1.01-1.11) |
| 63 | 1.43 (1.36-1.49) | 1.08 (1.07-1.08) | 1.85 (1.62-2.10) | 1.32 (1.26-1.37) | 2.31 (1.69-3.16) | 2.80 (2.56-3.07) | 0.63 (0.60-0.67) | 1.05 (1.00-1.09) |
| 64 | 1.48 (1.41-1.55) | 1.07 (1.06-1.07) | 1.76 (1.54-2.01) | 1.30 (1.25-1.35) | 2.50 (1.86-3.36) | 2.71 (2.47-2.98) | 0.62 (0.59-0.65) | 1.01 (0.97-1.06) |
| 65 | 1.48 (1.42-1.55) | 1.07 (1.06-1.07) | 1.74 (1.52-1.99) | 1.28 (1.23-1.34) | 2.63 (1.99-3.47) | 2.68 (2.44-2.94) | 0.63 (0.60-0.66) | 1.02 (0.98-1.07) |
| 66 | 1.47 (1.41-1.54) | 1.06 (1.06-1.07) | 1.69 (1.47-1.95) | 1.28 (1.23-1.33) | 2.72 (2.08-3.55) | 2.60 (2.37-2.86) | 0.62 (0.59-0.66) | 1.00 (0.96-1.05) |
| 67 | 1.44 (1.37-1.51) | 1.06 (1.06-1.07) | 1.71 (1.48-1.98) | 1.31 (1.25-1.36) | 2.50 (1.90-3.30) | 2.55 (2.32-2.80) | 0.64 (0.61-0.67) | 0.99 (0.94-1.03) |

| | | | | | | | | |
|---|---|---|---|---|---|---|---|---|
| 68 | 1.41 (1.34-1.48) | 1.05 (1.05-1.06) | 1.60 (1.37-1.88) | 1.28 (1.23-1.34) | 2.79 (2.14-3.65) | 2.29 (2.08-2.52) | 0.65 (0.62-0.69) | 0.97 (0.93-1.02) |
| 69 | 1.40 (1.33-1.47) | 1.05 (1.04-1.06) | 1.67 (1.40-1.98) | 1.26 (1.20-1.32) | 2.76 (2.08-3.66) | 2.14 (1.93-2.38) | 0.66 (0.63-0.70) | 0.97 (0.93-1.03) |
| 70 | 1.39 (1.30-1.48) | 1.05 (1.04-1.05) | 1.62 (1.28-2.04) | 1.20 (1.13-1.28) | 2.96 (2.10-4.17) | 1.99 (1.74-2.26) | 0.68 (0.63-0.73) | 1.01 (0.95-1.08) |

In the UK Biobank (UKB), we display the age-specific hazard ratio (HR) and 95% confidence interval (CI) per standardized increase in quantitative genomic and non-genomic risk factors, or for binary risk factors, the presence or absence of each.

**UKB**: UK Biobank; **PCE**: pooled cohort equations; **SBP**: Systolic Blood pressure; DM: Diabetes mellitus; **TC**: total cholesterol; **HDL**: high-density lipoprotein cholesterol.

**Supplementary Table 3.** Age-dependent proportion of variation explained by genomic and nongenomic risk factors in the FOS (N=3,588)

| Age (years) | PRS, PVE (95%CI) | Smoking, PVE (95% CI) | SBP, PVE (95% CI) | Total Cholesterol, PVE (95% CI) | Male sex, PVE (95% CI) | HDL-C, PVE (95% CI) |
|---|---|---|---|---|---|---|
| 18 | 0.19 (0.16, 0.22) | 0.06 (0.05, 0.08) | 0.04 (0.02, 0.05) | 0.00 (0.00, 0.00) | 0.04 (0.03, 0.05) | 0.01 (0.00, 0.01) |
| 19 | 0.11 (0.09, 0.13) | 0.06 (0.04, 0.07) | 0.05 (0.03, 0.06) | 0.00 (0.00, 0.01) | 0.05 (0.04, 0.06) | 0.02 (0.01, 0.03) |
| 20 | 0.14 (0.12, 0.16) | 0.03 (0.02, 0.05) | 0.05 (0.04, 0.07) | 0.01 (0.00, 0.01) | 0.03 (0.02, 0.05) | 0.01 (0.01, 0.02) |
| 21 | 0.09 (0.07, 0.11) | 0.02 (0.01, 0.03) | 0.06 (0.04, 0.07) | 0.00 (0.00, 0.00) | 0.03 (0.02, 0.05) | 0.00 (0.00, 0.00) |
| 22 | 0.06 (0.05, 0.08) | 0.02 (0.01, 0.03) | 0.06 (0.05, 0.08) | 0.00 (0.00, 0.00) | 0.04 (0.03, 0.05) | 0.00 (0.00, 0.00) |
| 23 | 0.06 (0.04, 0.07) | 0.02 (0.01, 0.03) | 0.06 (0.05, 0.08) | 0.00 (0.00, 0.00) | 0.05 (0.03, 0.06) | 0.01 (0.00, 0.01) |
| 24 | 0.06 (0.04, 0.07) | 0.03 (0.02, 0.04) | 0.06 (0.05, 0.08) | 0.00 (0.00, 0.01) | 0.04 (0.03, 0.06) | 0.01 (0.00, 0.01) |
| 25 | 0.04 (0.03, 0.05) | 0.02 (0.01, 0.03) | 0.04 (0.03, 0.05) | 0.00 (0.00, 0.00) | 0.04 (0.02, 0.05) | 0.02 (0.01, 0.03) |
| 26 | 0.02 (0.01, 0.03) | 0.01 (0.01, 0.02) | 0.02 (0.01, 0.03) | 0.00 (0.00, 0.00) | 0.04 (0.02, 0.05) | 0.01 (0.01, 0.02) |
| 27 | 0.02 (0.01, 0.03) | 0.02 (0.01, 0.03) | 0.03 (0.02, 0.04) | 0.00 (0.00, 0.00) | 0.04 (0.03, 0.05) | 0.02 (0.01, 0.03) |
| 28 | 0.03 (0.02, 0.05) | 0.02 (0.01, 0.03) | 0.03 (0.02, 0.04) | 0.00 (0.00, 0.01) | 0.03 (0.02, 0.04) | 0.03 (0.02, 0.04) |
| 29 | 0.02 (0.01, 0.03) | 0.02 (0.01, 0.02) | 0.02 (0.01, 0.03) | 0.00 (0.00, 0.01) | 0.02 (0.01, 0.03) | 0.02 (0.01, 0.03) |
| 30 | 0.02 (0.01, 0.03) | 0.02 (0.01, 0.02) | 0.02 (0.01, 0.03) | 0.01 (0.00, 0.01) | 0.03 (0.02, 0.04) | 0.03 (0.02, 0.04) |
| 31 | 0.02 (0.01, 0.03) | 0.01 (0.00, 0.02) | 0.03 (0.02, 0.04) | 0.01 (0.01, 0.02) | 0.03 (0.02, 0.04) | 0.04 (0.02, 0.05) |
| 32 | 0.02 (0.01, 0.03) | 0.01 (0.00, 0.02) | 0.03 (0.02, 0.04) | 0.01 (0.01, 0.02) | 0.02 (0.01, 0.03) | 0.03 (0.02, 0.04) |
| 33 | 0.02 (0.01, 0.03) | 0.01 (0.00, 0.02) | 0.03 (0.02, 0.04) | 0.01 (0.01, 0.02) | 0.02 (0.01, 0.03) | 0.03 (0.02, 0.04) |
| 34 | 0.02 (0.01, 0.03) | 0.01 (0.00, 0.02) | 0.03 (0.02, 0.04) | 0.02 (0.01, 0.02) | 0.03 (0.02, 0.04) | 0.03 (0.02, 0.04) |
| 35 | 0.03 (0.02, 0.04) | 0.01 (0.00, 0.01) | 0.02 (0.01, 0.03) | 0.02 (0.01, 0.02) | 0.02 (0.01, 0.03) | 0.03 (0.02, 0.04) |
| 36 | 0.03 (0.02, 0.04) | 0.01 (0.00, 0.02) | 0.02 (0.01, 0.03) | 0.02 (0.01, 0.03) | 0.03 (0.02, 0.04) | 0.04 (0.02, 0.05) |
| 37 | 0.03 (0.02, 0.04) | 0.01 (0.00, 0.02) | 0.02 (0.01, 0.03) | 0.02 (0.01, 0.03) | 0.02 (0.01, 0.03) | 0.03 (0.02, 0.04) |
| 38 | 0.03 (0.02, 0.04) | 0.01 (0.00, 0.02) | 0.02 (0.01, 0.03) | 0.02 (0.01, 0.03) | 0.03 (0.02, 0.04) | 0.03 (0.02, 0.04) |
| 39 | 0.03 (0.02, 0.04) | 0.01 (0.00, 0.02) | 0.02 (0.01, 0.03) | 0.03 (0.02, 0.04) | 0.03 (0.02, 0.04) | 0.04 (0.02, 0.05) |
| 40 | 0.03 (0.02, 0.04) | 0.01 (0.00, 0.02) | 0.02 (0.01, 0.03) | 0.02 (0.01, 0.03) | 0.03 (0.02, 0.04) | 0.03 (0.02, 0.05) |
| 41 | 0.03 (0.02, 0.04) | 0.01 (0.00, 0.02) | 0.02 (0.01, 0.03) | 0.02 (0.01, 0.03) | 0.03 (0.02, 0.04) | 0.04 (0.02, 0.05) |
| 42 | 0.03 (0.02, 0.04) | 0.01 (0.00, 0.02) | 0.02 (0.01, 0.03) | 0.03 (0.02, 0.04) | 0.03 (0.02, 0.04) | 0.04 (0.03, 0.05) |
| 43 | 0.03 (0.02, 0.04) | 0.01 (0.00, 0.01) | 0.02 (0.01, 0.03) | 0.03 (0.02, 0.04) | 0.03 (0.02, 0.04) | 0.04 (0.03, 0.05) |

| | | | | | | |
|---|---|---|---|---|---|---|
| 44 | 0.03 (0.02, 0.04) | 0.01 (0.00, 0.01) | 0.02 (0.01, 0.03) | 0.03 (0.02, 0.04) | 0.03 (0.02, 0.05) | 0.04 (0.03, 0.05) |
| 45 | 0.03 (0.02, 0.05) | 0.01 (0.00, 0.01) | 0.02 (0.01, 0.03) | 0.03 (0.02, 0.04) | 0.03 (0.02, 0.04) | 0.04 (0.03, 0.06) |
| 46 | 0.04 (0.02, 0.05) | 0.01 (0.00, 0.01) | 0.02 (0.01, 0.03) | 0.04 (0.03, 0.05) | 0.03 (0.02, 0.04) | 0.04 (0.03, 0.05) |
| 47 | 0.03 (0.02, 0.05) | 0.01 (0.00, 0.01) | 0.03 (0.02, 0.04) | 0.04 (0.03, 0.05) | 0.03 (0.02, 0.04) | 0.03 (0.02, 0.05) |
| 48 | 0.03 (0.02, 0.04) | 0.01 (0.00, 0.01) | 0.03 (0.02, 0.04) | 0.04 (0.03, 0.06) | 0.03 (0.02, 0.04) | 0.03 (0.02, 0.05) |
| 49 | 0.03 (0.02, 0.04) | 0.01 (0.00, 0.01) | 0.03 (0.02, 0.04) | 0.04 (0.03, 0.06) | 0.03 (0.02, 0.04) | 0.04 (0.02, 0.05) |
| 50 | 0.03 (0.02, 0.04) | 0.01 (0.00, 0.01) | 0.03 (0.02, 0.04) | 0.05 (0.03, 0.06) | 0.03 (0.02, 0.04) | 0.03 (0.02, 0.04) |
| 51 | 0.03 (0.02, 0.04) | 0.01 (0.00, 0.01) | 0.03 (0.02, 0.04) | 0.04 (0.03, 0.06) | 0.03 (0.02, 0.04) | 0.03 (0.02, 0.04) |
| 52 | 0.03 (0.02, 0.04) | 0.01 (0.00, 0.01) | 0.03 (0.02, 0.04) | 0.05 (0.03, 0.06) | 0.03 (0.02, 0.04) | 0.03 (0.02, 0.04) |
| 53 | 0.03 (0.02, 0.04) | 0.01 (0.00, 0.01) | 0.03 (0.02, 0.04) | 0.04 (0.03, 0.06) | 0.03 (0.02, 0.04) | 0.03 (0.02, 0.04) |
| 54 | 0.03 (0.02, 0.04) | 0.01 (0.00, 0.01) | 0.03 (0.02, 0.04) | 0.04 (0.03, 0.06) | 0.03 (0.02, 0.04) | 0.03 (0.02, 0.04) |
| 55 | 0.03 (0.02, 0.04) | 0.01 (0.00, 0.01) | 0.03 (0.02, 0.04) | 0.04 (0.03, 0.06) | 0.03 (0.02, 0.04) | 0.03 (0.02, 0.04) |
| 56 | 0.03 (0.02, 0.04) | 0.01 (0.00, 0.01) | 0.03 (0.02, 0.04) | 0.04 (0.03, 0.06) | 0.03 (0.02, 0.04) | 0.03 (0.02, 0.04) |
| 57 | 0.03 (0.02, 0.04) | 0.01 (0.00, 0.01) | 0.03 (0.02, 0.04) | 0.04 (0.03, 0.06) | 0.03 (0.02, 0.04) | 0.03 (0.02, 0.04) |

In the Framingham offspring study (FOS), we display the age-specific proportion of variation explained for individuals up to and including the age of consideration. 95% CI calculated according to Owen et al (2003)[1]. **FOS**: Framingham Offspring Study; **TC**: Total Cholesterol, **SBP**: Systolic Blood pressure; **HDL**: High density lipoprotein cholesterol, **PRS**: Polygenic Risk Score.

**Supplementary Table 4: Age-dependent proportion of variation explained by genomic and nongenomic risk factors in the UKB (N=327,837)**

| Age (years) | PRS, PVE (95%CI) | Smoking, PVE (95% CI) | SBP, PVE (95%CI) | DM, PVE (95%CI) | Male sex, PVE (95% CI) | HDL, PVE (95%CI) | PCE-SD,PVE (95%CI) |
|---|---|---|---|---|---|---|---|
| 41 | 0.059 (0.057, 0.061) | 0.027 (0.026, 0.028) | 0.009 (0.008, 0.010) | 0.001 (0.001, 0.001) | 0.025 (0.024, 0.026) | 0.045 (0.044, 0.046) | 0.062 (0.060, 0.063) |
| 42 | 0.042 (0.041, 0.043) | 0.027 (0.026, 0.028) | 0.014 (0.013, 0.015) | 0.000 (0.000, 0.000) | 0.034 (0.031, 0.037) | 0.040 (0.039, 0.041) | 0.056 (0.055, 0.058) |
| 43 | 0.028 (0.027, 0.029) | 0.019 (0.018, 0.020) | 0.018 (0.017, 0.019) | 0.000 (0.000, 0.000) | 0.031 (0.028, 0.034) | 0.036 (0.035, 0.037) | 0.055 (0.054, 0.057) |
| 44 | 0.028 (0.027, 0.029) | 0.023 (0.022, 0.024) | 0.019 (0.018, 0.020) | 0.000 (0.000, 0.000) | 0.033 (0.030, 0.036) | 0.036 (0.035, 0.037) | 0.064 (0.062, 0.066) |
| 45 | 0.026 (0.025, 0.027) | 0.023 (0.022, 0.024) | 0.024 (0.023, 0.025) | 0.000 (0.000, 0.000) | 0.037 (0.034, 0.040) | 0.034 (0.033, 0.035) | 0.072 (0.071, 0.074) |
| 46 | 0.027 (0.026, 0.028) | 0.021 (0.020, 0.022) | 0.027 (0.026, 0.028) | 0.001 (0.001, 0.001) | 0.038 (0.035, 0.041) | 0.032 (0.031, 0.033) | 0.072 (0.071, 0.074) |
| 47 | 0.025 (0.024, 0.026) | 0.020 (0.019, 0.021) | 0.026 (0.025, 0.027) | 0.000 (0.000, 0.000) | 0.037 (0.034, 0.040) | 0.029 (0.028, 0.030) | 0.074 (0.072, 0.076) |
| 48 | 0.026 (0.025, 0.027) | 0.019 (0.018, 0.020) | 0.026 (0.025, 0.027) | 0.001 (0.001, 0.001) | 0.035 (0.032, 0.038) | 0.030 (0.029, 0.031) | 0.074 (0.072, 0.076) |
| 49 | 0.027 (0.026, 0.028) | 0.017 (0.016, 0.018) | 0.027 (0.026, 0.028) | 0.001 (0.001, 0.001) | 0.035 (0.032, 0.038) | 0.030 (0.029, 0.031) | 0.073 (0.071, 0.075) |
| 50 | 0.026 (0.025, 0.027) | 0.017 (0.016, 0.018) | 0.026 (0.025, 0.027) | 0.001 (0.001, 0.001) | 0.035 (0.032, 0.038) | 0.028 (0.027, 0.029) | 0.071 (0.069, 0.073) |
| 51 | 0.026 (0.025, 0.027) | 0.016 (0.015, 0.017) | 0.027 (0.026, 0.028) | 0.001 (0.001, 0.001) | 0.035 (0.032, 0.038) | 0.027 (0.026, 0.028) | 0.072 (0.070, 0.073) |
| 52 | 0.026 (0.025, 0.027) | 0.014 (0.013, 0.015) | 0.025 (0.024, 0.026) | 0.001 (0.001, 0.001) | 0.036 (0.033, 0.039) | 0.026 (0.025, 0.027) | 0.069 (0.067, 0.071) |
| 53 | 0.024 (0.023, 0.025) | 0.014 (0.013, 0.015) | 0.025 (0.024, 0.026) | 0.001 (0.001, 0.001) | 0.036 (0.033, 0.039) | 0.025 (0.024, 0.026) | 0.069 (0.067, 0.071) |
| 54 | 0.023 (0.022, 0.024) | 0.014 (0.013, 0.015) | 0.024 (0.023, 0.025) | 0.001 (0.001, 0.001) | 0.036 (0.033, 0.039) | 0.023 (0.022, 0.024) | 0.070 (0.068, 0.071) |
| 55 | 0.024 (0.023, 0.025) | 0.013 (0.012, 0.014) | 0.023 (0.022, 0.024) | 0.001 (0.001, 0.001) | 0.036 (0.033, 0.039) | 0.024 (0.023, 0.025) | 0.069 (0.067, 0.071) |
| 56 | 0.024 (0.023, 0.025) | 0.013 (0.012, 0.014) | 0.023 (0.022, 0.024) | 0.001 (0.001, 0.001) | 0.036 (0.033, 0.039) | 0.024 (0.023, 0.025) | 0.069 (0.067, 0.070) |
| 57 | 0.023 (0.022, 0.024) | 0.012 (0.011, 0.013) | 0.023 (0.022, 0.024) | 0.001 (0.001, 0.001) | 0.036 (0.033, 0.039) | 0.024 (0.023, 0.025) | 0.070 (0.068, 0.071) |
| 58 | 0.023 (0.022, 0.024) | 0.011 (0.010, 0.012) | 0.023 (0.022, 0.024) | 0.001 (0.001, 0.001) | 0.035 (0.032, 0.038) | 0.024 (0.023, 0.025) | 0.070 (0.068, 0.071) |

| | | | | | | | |
|---|---|---|---|---|---|---|---|
| 59 | 0.023 (0.022, 0.024) | 0.011 (0.010, 0.012) | 0.023 (0.022, 0.024) | 0.001 (0.001, 0.001) | 0.036 (0.033, 0.039) | 0.025 (0.024, 0.026) | 0.070 (0.068, 0.071) |
| 60 | 0.022 (0.021, 0.023) | 0.010 (0.009, 0.011) | 0.024 (0.023, 0.025) | 0.001 (0.001, 0.001) | 0.034 (0.031, 0.037) | 0.025 (0.024, 0.026) | 0.070 (0.068, 0.072) |
| 61 | 0.020 (0.019, 0.021) | 0.009 (0.008, 0.010) | 0.024 (0.023, 0.025) | 0.001 (0.001, 0.001) | 0.034 (0.031, 0.037) | 0.025 (0.024, 0.026) | 0.070 (0.069, 0.072) |
| 62 | 0.019 (0.018, 0.020) | 0.009 (0.008, 0.010) | 0.023 (0.022, 0.024) | 0.001 (0.001, 0.001) | 0.033 (0.030, 0.036) | 0.024 (0.023, 0.025) | 0.069 (0.067, 0.070) |
| 63 | 0.019 (0.018, 0.020) | 0.008 (0.007, 0.009) | 0.024 (0.023, 0.025) | 0.001 (0.001, 0.001) | 0.033 (0.030, 0.036) | 0.023 (0.022, 0.024) | 0.068 (0.066, 0.070) |
| 64 | 0.018 (0.017, 0.019) | 0.007 (0.006, 0.008) | 0.024 (0.023, 0.025) | 0.001 (0.001, 0.001) | 0.032 (0.029, 0.035) | 0.022 (0.021, 0.023) | 0.067 (0.065, 0.069) |
| 65 | 0.018 (0.017, 0.019) | 0.007 (0.006, 0.008) | 0.023 (0.022, 0.024) | 0.002 (0.002, 0.002) | 0.032 (0.029, 0.035) | 0.022 (0.021, 0.023) | 0.066 (0.064, 0.067) |
| 66 | 0.018 (0.017, 0.019) | 0.006 (0.005, 0.007) | 0.024 (0.023, 0.025) | 0.002 (0.002, 0.002) | 0.032 (0.029, 0.035) | 0.022 (0.021, 0.023) | 0.066 (0.065, 0.068) |
| 67 | 0.018 (0.017, 0.019) | 0.006 (0.005, 0.007) | 0.024 (0.023, 0.025) | 0.002 (0.002, 0.002) | 0.031 (0.028, 0.034) | 0.021 (0.020, 0.022) | 0.066 (0.064, 0.068) |
| 68 | 0.017 (0.016, 0.018) | 0.005 (0.005, 0.005) | 0.025 (0.024, 0.026) | 0.002 (0.002, 0.002) | 0.031 (0.028, 0.034) | 0.021 (0.020, 0.022) | 0.066 (0.065, 0.068) |
| 69 | 0.017 (0.016, 0.018) | 0.005 (0.005, 0.005) | 0.025 (0.024, 0.026) | 0.002 (0.002, 0.002) | 0.030 (0.027, 0.033) | 0.021 (0.020, 0.022) | 0.066 (0.064, 0.068) |
| 70 | 0.017 (0.016, 0.018) | 0.005 (0.005, 0.005) | 0.025 (0.024, 0.026) | 0.002 (0.002, 0.002) | 0.029 (0.026, 0.032) | 0.021 (0.020, 0.022) | 0.065 (0.064, 0.067) |

In the UK Biobank, we display the age-specific proportion of variation explained for individuals up to and including the age of consideration. 95% CI calculated according to Owen et al (2003)[1]. **FOS**: Framingham Offspring Study; **TC**: Total Cholesterol; **SBP**; Systolic Blood pressure; **HDL**: High density lipoprotein cholesterol, **PRS**: Polygenic Risk Score, **PCE**: Pooled cohort equations; **PCE-SD:** Pooled cohort equations scaled by their standard deviation.

**Supplementary Table 5: Translation of age-specific PCE clinical risk quintiles to predicted 10-year ASCVD risk (N=327,837)**

| Age group | Age-specific PCE Category | Predicted 10-year ASCVD-Risk (%) |
|---|---|---|
| <55 | Low (bottom quintile) | < 1.0 |
| <55 | Intermediate (middle three quintiles) | 1.0-5.1 |
| <55 | High (top quintile) | >5.1 |
| 55-65 | Low (bottom quintile) | <3.6 |
| 55-65 | Intermediate (middle three quintiles) | 3.6-13.6 |
| 55-65 | High (top quintile) | >13.6 |
| >65 | Low (bottom quintile) | <8.7 |
| >65 | Intermediate (middle three quintiles) | 8.7-23.2 |
| >65 | High (top quintile) | >23.2 |

Because the distribution of PCE 10-year ASCVD risk estimates is very different by age groups making direct comparisons challenging, we report the10-year predicted ASCVD risk for individuals by age-specific quantiles. For each age we compute the age-specific PCE quantile and the associated predicted 10-year ASCVD risk. **PCE:** Pooled Cohort Equations, **ASCVD:** Atherosclerotic Cardiovascular Disease.

**Supplementary Table 6: Proportion of CAD events predicted by genomic and nongenomic risk by age of estimation in the UKB (N=327,837)**

| Age Group | Risk Stratification | Proportion Detected | Lower 95% CI | Upper 95% CI |
|---|---|---|---|---|
| 40-45 | High PRS | 0.32 | 0.32 | 0.33 |
| 45-50 | High PRS | 0.29 | 0.29 | 0.29 |
| 50-55 | High PRS | 0.17 | 0.17 | 0.17 |
| 55-60 | High PRS | 0.12 | 0.12 | 0.12 |
| 60-65 | High PRS | 0.06 | 0.06 | 0.06 |
| 65-70 | High PRS | 0.01 | 0.01 | 0.01 |
| 70-75 | High PRS | 0.00 | 0.00 | 0.00 |
| 40-45 | Both | 0.09 | 0.09 | 0.09 |
| 45-50 | Both | 0.11 | 0.10 | 0.11 |
| 50-55 | Both | 0.17 | 0.17 | 0.18 |
| 55-60 | Both | 0.23 | 0.23 | 0.23 |
| 60-65 | Both | 0.24 | 0.24 | 0.24 |
| 65-70 | Both | 0.27 | 0.27 | 0.27 |
| 70-75 | Both | 0.23 | 0.21 | 0.24 |
| 40-45 | High PCE | 0.09 | 0.09 | 0.09 |
| 45-50 | High PCE | 0.15 | 0.15 | 0.15 |
| 50-55 | High PCE | 0.27 | 0.27 | 0.27 |
| 55-60 | High PCE | 0.44 | 0.44 | 0.44 |
| 60-65 | High PCE | 0.55 | 0.55 | 0.55 |
| 65-70 | High PCE | 0.68 | 0.68 | 0.69 |
| 70-75 | High PCE | 0.77 | 0.76 | 0.79 |

Within five-year age strata, we identify the proportion of cases in that age group who go on to develop CAD and were correctly stratified at enrollment by genomic risk stratification (high PRS), clinical risk stratification (high PCE), and both. **UKB**: UK Biobank, **PRS**: Polygenic Risk Score, **PCE**: Pooled Cohort Equations.

**Supplementary Table 7: Characteristics of study participants by genomic risk category in the UKB (N=327,837)**

| | Low | Intermediate | High | Overall |
|---|---|---|---|---|
| | (N=65696) | (N=196750) | (N=65391) | (N=327837) |
| Age at risk estimation, mean (SD), years | 56.1 (8.14) | 56.2 (8.07) | 55.7 (8.05) | 56.1 (8.08) |
| Female, n (%) | 36939 (56.2%) | 111556 (56.7%) | 38012 (58.1%) | 186507 (56.9%) |
| White, n (%) | 51294 (78.1%) | 168637 (85.7%) | 54996 (84.1%) | 274927 (83.9%) |
| Incident CAD, n (%) | 1290 (2.0%) | 6282 (3.2%) | 3618 (5.5%) | 11190 (3.4%) |
| Follow-up period, median (IQR) | 11.7 (1.78) | 11.7 (1.92) | 11.6 (2.15) | 11.7 (1.94) |
| Diabetes Mellitus, n (%) | 541 (0.8%) | 1375 (0.7%) | 497 (0.8%) | 2413 (0.7%) |
| Current Smoking, n (%) | 6730 (10.2%) | 20181 (10.3%) | 6958 (10.6%) | 33869 (10.3%) |
| Total cholesterol, mean (SD), mg/dL | 222 (40.7) | 229 (41.1) | 234 (42.1) | 229 (41.4) |
| HDL Cholesterol, mean (SD), mg/dL | 57.6 (14.9) | 57.1 (14.7) | 56.8 (14.7) | 57.2 (14.8) |
| LDL cholesterol, mean (SD), mg/dL | 139 (31.4) | 144 (31.7) | 149 (32.4) | 144 (31.9) |
| Triglycerides, mean (SD), mg/dL | 145 (85.9) | 151 (87.9) | 153 (89.5) | 150 (87.9) |
| Diastolic blood pressure, mean (SD), mmHg | 82.3 (11.2) | 82.6 (11.1) | 83.1 (11.0) | 82.7 (11.1) |
| Systolic blood pressure, mean (SD), mmHg | 139 (20.3) | 140 (20.4) | 141 (20.4) | 140 (20.4) |
| Taking antihypertensive medication, n (%) | 8073 (12.3%) | 24279 (12.3%) | 8736 (13.4%) | 41088 (12.5%) |
| PCE 10-year ASCVD risk category | | | | |
| Low or borderline (<7.5%), n (%) | 42166 (64.2%) | 123405 (62.7%) | 41579 (63.6%) | 207150 (63.2%) |
| Intermediate (≥7.5 to <20%), n (%) | 18892 (28.8%) | 58686 (29.8%) | 19197 (29.4%) | 96775 (29.5%) |
| High (≥20%), n (%) | 4638 (7.1%) | 14659 (7.5%) | 4615 (7.1%) | 23912 (7.3%) |

The table highlights the clinical risk factors for individuals in the UKB as stratified by genomic risk – low (PRS in the bottom quintile), intermediate (PRS in the middle three quintiles), and high (PRS in the top quintile). UKB: UK Biobank, PRS: Polygenic Risk Score.

**Supplementary Table 8: Characteristics of study participants by clinical risk category in the UKB (N=327,837)**

| | Low | Intermediate | High | Overall |
|---|---|---|---|---|
| | (N=65696) | (N=196750) | (N=65391) | (N=327837) |
| Age at risk estimation, mean (SD), years | 52.5 (7.08) | 61.4 (5.83) | 65.4 (3.79) | 56.1 (8.08) |
| Female, n (%) | 152147 (73.4%) | 33659 (34.8%) | 701 (2.9%) | 186507 (56.9%) |
| White, n (%) | 169969 (82.1%) | 83889 (86.7%) | 21069 (88.1%) | 274927 (83.9%) |
| Incident CAD, n (%) | 3264 (1.6%) | 5270 (5.4%) | 2656 (11.1%) | 11190 (3.4%) |
| Follow-up period, median (IQR) | 12.1 (11.39-12.73) | 12.0 (11.14 -12.65) | 11.7 (10.86 13.96) | 12.0 (11.31-12.69) |
| Diabetes Mellitus, n (%) | 537 (0.3%) | 813 (0.8%) | 1063 (4.4%) | 2413 (0.7%) |
| Current Smoking, n (%) | 13871 (6.7%) | 14798 (15.3%) | 5200 (21.7%) | 33869 (10.3%) |
| Total cholesterol, mean (SD), mg/dL | 224 (40.3) | 237 (41.9) | 235 (42.4) | 229 (41.4) |
| HDL Cholesterol, mean (SD), mg/dL | 59.7 (14.8) | 54.0 (13.7) | 48.0 (11.6) | 57.2 (14.8) |
| LDL cholesterol, mean (SD), mg/dL | 139 (30.8) | 152 (31.8) | 154 (33.1) | 144 (31.9) |
| Triglycerides, mean (SD), mg/dL | 145 (85.9) | 151 (87.9) | 153 (89.5) | 150 (87.9) |
| Diastolic blood pressure, mean (SD), mmHg | 82.3 (11.2) | 82.6 (11.1) | 83.1 (11.0) | 82.7 (11.1) |
| Systolic blood pressure, mean (SD), mmHg | 133 (17.1) | 148 (18.4) | 166 (19.1) | 140 (20.4) |
| Taking antihypertensive medication, n (%) | 16129 (7.8%) | 15804 (16.3%) | 9155 (38.3%) | 41088 (12.5%) |
| Genomic Risk Category (PRS) | | | | |
| Low ( PRS <20%), n (%) | 42166 (20.4%) | 18892 (19.5%) | 4638 (19.4%) | 65696 (20.0%) |
| Intermediate (PRS ≥20 to <80%), n (%) | 123405 (59.6%) | 58686 (60.6%) | 14659 (61.3%) | 196750 (60.0%) |
| High (PRS ≥80%), n (%) | 41579 (20.1%) | 19197 (19.8%) | 4615 (19.3%) | 65391 (19.9%) |

The table highlights the clinical risk factors for individuals in the UKB as stratified by clinical risk – low or borderline (PCE 10-year ASCVD risk <7.5%), intermediate (PCE 10-year ASCVD risk 7.5-20%) and high (PCE 10-year ASCVD risk >20%). UKB: UK Biobank, PCE: Pooled Cohort Equations, ASCVD: Atherosclerotic Cardiovascular Disease, PRS: Polygenic Risk Score

**Supplementary Table 9: Mean age of CAD event by genomic or clinical risk decile in the UKB (N=327,837)**

| | Genomic Risk (PRS) | | | Clinical Risk (PCE 10-year ASCVD) | | |
|---|---|---|---|---|---|---|
| Percentile | Number of Events | Mean Age of Event (years) | SD (years) | Number of Events | Mean Age of Event (years) | SD (years) |
| 10 | 646 | 67.17 | 7.70 | 110 | 55.19 | 5.08 |
| 20 | 785 | 67.28 | 7.47 | 275 | 57.62 | 6.06 |
| 30 | 837 | 67.08 | 7.64 | 446 | 58.94 | 6.96 |
| 40 | 918 | 67.34 | 7.46 | 657 | 61.09 | 7.53 |
| 50 | 1028 | 66.75 | 7.65 | 869 | 62.94 | 7.49 |
| 60 | 1126 | 66.52 | 7.53 | 1180 | 64.46 | 7.51 |
| 70 | 1256 | 66.57 | 7.56 | 1451 | 66.34 | 7.26 |
| 80 | 1424 | 65.79 | 7.92 | 2026 | 67.56 | 6.75 |
| 90 | 1689 | 65.58 | 7.77 | 2972 | 70.06 | 5.69 |
| 100 | 1217 | 64.50 | 8.02 | 1100 | 68.87 | 7.37 |

Number and mean age of event by overall PRS and PCE decile in the UKB. PRS: Polygenic Risk Score, **PCE:** Pooled Cohort Equation, **UKB**: UK Biobank; **SD**: standard deviation., ASCVD: Atherosclerotic Cardiovascular Disease

**Supplementary Table 10: Discrimination of clinical vs. combined clinical-genomic risk models by age in the UKB (N=327,837)**

| Age | Model | AUC | SE | upper | lower |
|---|---|---|---|---|---|
| 40-45 | Clinical | 0.55 | 0.01 | 0.56 | 0.54 |
| 45-50 | Clinical | 0.60 | 0.01 | 0.61 | 0.59 |
| 50-55 | Clinical | 0.64 | 0.01 | 0.65 | 0.63 |
| 55-60 | Clinical | 0.66 | 0.01 | 0.67 | 0.65 |
| 60-65 | Clinical | 0.65 | 0.01 | 0.66 | 0.65 |
| 65-70 | Clinical | 0.62 | 0.01 | 0.63 | 0.62 |
| 70-75 | Clinical | 0.56 | 0.04 | 0.60 | 0.52 |
| 40-45 | Combined | 0.64 | 0.02 | 0.66 | 0.62 |
| 45-50 | Combined | 0.68 | 0.01 | 0.69 | 0.67 |
| 50-55 | Combined | 0.69 | 0.01 | 0.70 | 0.68 |
| 55-60 | Combined | 0.70 | 0.01 | 0.71 | 0.69 |
| 60-65 | Combined | 0.68 | 0.01 | 0.69 | 0.68 |
| 65-70 | Combined | 0.65 | 0.01 | 0.66 | 0.65 |
| 70-75 | Combined | 0.60 | 0.04 | 0.64 | 0.55 |

We report the Area Under the Receiver Operator Curve (AUC) for a model using genomic (PRS), clinical (PCE) or a combined metric to predict incident coronary artery disease. To isolate effect of each score, we use a logistic regression that uses the ordinal level of genomic (<20%, 20-80%, or >80%) or clinical (<7.5%, 7.5%-20%,>20% 10-year risk) in predicting incident coronary disease. For each ROC metric, we divide into a training and testing data set and report the AUC of the model developed in the training data on predicting incident coronary disease in the testing set. **ROC**: Receiver Operator Curve, **AUC**: Area Under Curve.

**Supplementary Table 11: Net reclassification of CAD events and non-events with the CAD PRS by age in the UK Biobank (N=327,837)**

| Ages | Variable | Value | Standard Error |
|---|---|---|---|
| <50 | Net Reclassification Event | 0.1612 | 0.0012 |
| <55 | Net Reclassification Event | 0.0341 | 0.0005 |
| <60 | Net Reclassification Event | -0.1078 | 0.0007 |
| <65 | Net Reclassification Event | -0.2507 | 0.0008 |
| <70 | Net Reclassification Event | -0.3532 | 0.0008 |
| <75 | Net Reclassification Event | -0.3559 | 0.0008 |
| <50 | Net Reclassification Non-event | -0.1550 | 0.0012 |
| <55 | Net Reclassification Non-event | -0.1097 | 0.0008 |
| <60 | Net Reclassification Non-event | -0.0347 | 0.0004 |
| <65 | Net Reclassification Non-event | 0.0600 | 0.0005 |
| <70 | Net Reclassification Non-event | 0.1491 | 0.0006 |
| <75 | Net Reclassification Non-event | 0.1514 | 0.0006 |

We report the net reclassification of events and non-events for individuals in the UK Biobank comparing the use of PRS alone over PCE alone for individuals up to the age in question. Net Reclassification Event is defined as Cases Correctly Reclassified by PRS versus PCE divided by the total number of cases. Net Reclassification Non-Event defined as Controls correctly Reclassified by PRS versus PCE divided by total number of controls. **PRS**: Polygenic Risk Score, **PCE**: Pooled Cohort Equations

**Supplementary Table 12: Proportion of individuals by age in each predicted 10-year ASCVD risk category in the UKB (327,837).**

| **Age** | Predicted 10-year ASCVD risk (%) | Proportional representation | 95% lower CI | 95% upper CI |
|---|---|---|---|---|
| 40-45 | <7.5 | 0.96 | 0.96 | 0.97 |
| 45-50 | <7.5 | 0.93 | 0.93 | 0.93 |
| 50-55 | <7.5 | 0.82 | 0.82 | 0.83 |
| 55-60 | <7.5 | 0.64 | 0.64 | 0.65 |
| 60-65 | <7.5 | 0.46 | 0.46 | 0.46 |
| 65-70 | <7.5 | 0.12 | 0.12 | 0.12 |
| 70-75 | <7.5 | 0.01 | 0.01 | 0.01 |
| 40-45 | 7.5-20 | 0.03 | 0.03 | 0.03 |
| 45-50 | 7.5-20 | 0.06 | 0.06 | 0.06 |
| 50-55 | 7.5-20 | 0.17 | 0.17 | 0.17 |
| 55-60 | 7.5-20 | 0.33 | 0.33 | 0.33 |
| 60-65 | 7.5-20 | 0.45 | 0.44 | 0.45 |
| 65-70 | 7.5-20 | 0.59 | 0.59 | 0.59 |
| 70-75 | 7.5-20 | 0.60 | 0.58 | 0.61 |
| 40-45 | >20 | 0.00 | 0.00 | 0.00 |
| 45-50 | >20 | 0.00 | 0.00 | 0.00 |
| 50-55 | >20 | 0.01 | 0.01 | 0.01 |
| 55-60 | >20 | 0.02 | 0.02 | 0.03 |
| 60-65 | >20 | 0.10 | 0.09 | 0.10 |
| 65-70 | >20 | 0.29 | 0.29 | 0.29 |
| 70-75 | >20 | 0.39 | 0.38 | 0.41 |

We display the proportion of individuals in each age interval with corresponding predicted PCE 10-year ASCVD risk. **PCE**: Pooled Cohort Equations, **ASCVD**: Atherosclerotic Cardiovascular Disease

### Supplementary Methods

#### Demonstrating Resolution of Non-proportional Hazards using Age Binning

As detailed in Methods, the effect of the covariates contained in ***X*** on the instantaneous hazard experienced by an individual is assumed to remain constant over time. Schoenfeld residuals reflect[2, 3] the observed minus the expected values of the covariates at each failure time, and we report the p-value testing of these residuals are independent of time under our model compared to a model agnostic to enrollment time (Supplementary Figure 3). We demonstrate that the proportional hazards assumption is resolved using age-stratified models (Supplementary Figure 3a). We use locally estimated scatterplot smoothing (LOESS) through the response variable average and display the SEM of the line of best fit and CI for the HR. The loess[4] smoother uses locally estimated weighted regression to weight each interval estimate by the tricube distance of nearby points, thereby borrowing information from nearby estimates to improve the precision of our estimates for intervals in which the case count may have been low. This was particularly important for the FOS cohort.

#### Computing Age-Specific Hazard Ratio and Proportion of Variation Explained

Let X= ($X_{i1}, \ldots, X_{ip}$) be the realized values of the covariates for subject *i*, the hazard function for the Cox proportional hazards model has the form

**Equation 1**

$$\lambda\,(t|X_i) = \lambda_{\,0}(t)\, \exp\left(\beta_{\,1}X_{i1} + \cdots\ + \beta_{\,p}X_{ip}\right)$$
$$= \lambda_0(t)\exp(X_i\beta)$$

where $(X_i\beta)$ represents the vector of covariates for individual i. This expression gives the hazard function at time t for subject i with covariate vector (explanatory variables) $X_i$. The hazard ratio for two subjects **i** and **j** with different levels of covariates is thus of the form:

**Equation 2**

$$\mathrm{HR} = \frac{\lambda_0(t)\exp(\beta_1 X_i)}{\lambda_0(t)\exp(\beta_1 X_j)}$$

$$= \frac{\exp(\beta_1 X_i)}{\exp(\beta_1 X_j)}$$

A fundamental assumption above is that while the baseline hazard $\lambda_0(t)$ is dependent on time (t), the hazard ratio (HR) between two subjects *i* and *j* with different covariate levels is independent of time and thus remains proportional over the time period considered (Equation 2).

For binary variables, we reported the HR with respect to the presence (over the absence) of a given risk factor. PRS, LDL-C, HDL-C, total cholesterol, body mass index, and systolic blood pressure were standardized such that the mean and standard deviation were 0 and 1 respectively.

For PRS, we rescale to a normal distribution (0,1) for all individuals who meet the study criteria. For the PCE, the ten-year risk estimates as calculated by the PCE above were normalized to a (0,1) distribution and HR per SD[5] was reported. For proportion of variation explained calculation, we used McFadden's Pseudo $R^2$ as: 1-deviance/null Deviance of the logistic regression for individuals less than or equal to the age of consideration using a probit link function.

$$\log\frac{p}{1-p} = \beta_0 + \beta_1 x_1$$

$$R^2_{McF} = 1 - \frac{lnL_m}{lnL_0}$$

**Calculating Net Reclassification**

For net reclassification indices, at each age of consideration, we defined $NRI_{event}$ as the net proportion of cases correctly reclassified by genomic risk:

$$NRI_{event}$$

$$\frac{Genomic\ Risk > 0.80\ \cap PCE < 7.5 \cap CAD - Genomic\ Risk < 0.80\ \cap PCE > 7.5 \cap CAD}{Develop\ CAD}$$

We defined $NRI_{non\text{-}event}$ as the net proportion of controls correctly reclassified by genomic risk:

$$NRI_{non-event}$$

$$\frac{Genomic\ Risk < 0.80\ \cap PCE > 7.5 \cap No\ CAD - Genomic\ Risk > 0.80\ \cap PCE < 7.5 \cap No\ CAD}{Does\ not\ develop\ CAD}$$

**Delta AUC**

We use DeLong's test for delta AUC and the test proposed by Pepe *et al* to assess for difference in sensitivity at a given level of specificity.[4,5] Net reclassification of events at different ages of consideration were calculated (Supplemental Methods). Statistical analyses were performed using R software, version 4.2.1 (R Project for Statistical Computing).
